# Digital health app data reveals an effect of ovarian hormones on long COVID and myalgic encephalomyelitis symptoms

**DOI:** 10.1101/2025.01.24.25321092

**Authors:** Abigail Goodship, Rory Preston, Joseph T Hicks, Natalie Getreu, Helen O’Neill, Harry Leeming, Christian Morgenstern, Victoria Male

## Abstract

**Background:** Long COVID and myalgic encephalomyelitis/chronic fatigue syndrome (ME/CFS) disproportionately affect females, suggesting modulation by sex hormones. We investigated whether symptom severity is influenced by changes in sex hormones over the menstrual cycle, or by hormonal contraception.

**Methods:** We carried out a retrospective analysis of menstrual and symptom data, prospectively collected via the Visible app from individuals with long COVID, ME/CFS, or both, who had regular menstrual cycles (7th September 2022-6th March 2024). We examined associations between symptom severity, menstrual cycle phase and contraception type using mixed-effects models. Prospectively collected menstrual and symptom data from users of the Hertility menstrual cycle tracking app (7^th^ September 2022-28^th^ September 2025) were used as a population comparison cohort.

**Findings:** 948 users were included in the disease cohort; 100% of users were female, 92.6% identified as women, 6.4% as non-binary, 0.6% preferred not to say and 0.3% self-described. Crashes (sudden and severe worsening of symptoms following exertion) were significantly more frequent during menstruation than other phases, most notably compared with the late luteal phase (odds ratio (OR) = 0·888, 95% confidence interval (CI): 0·838-0·941). Users of combined hormonal contraception (n=70) had reduced crash incidence (OR = 0·548, 95% CI: 0·350–0·856) and overall symptom score (incidence rate ratio = 0·827, 95% CI: 0·690–0·992) compared to those not using contraception (n=786). Fatigue, brain fog, and headaches showed phase-specific variation in both disease and population comparison cohorts.

**Interpretation:** Menstruation is associated with increased crash incidence and worsened symptoms in long COVID and ME/CFS. Users of combined hormonal contraception report reduced crash incidence and lower symptom burden, suggesting a modulatory role of ovarian hormones. Analysis of the population comparison cohort indicates that some cyclical symptom variation may reflect normal menstrual-related fluctuations, albeit that symptoms are less frequently reported in the general population. Our models adjusted for factors including individual, disease type, age and gravidity, however, residual confounding accounting for differences in disease severity between contraceptive users and non-users cannot be excluded. Nevertheless, these findings could empower menstruating people living with long COVID and ME/CFS to anticipate cyclical changes in symptoms and plan their activities accordingly and could also inform their use of contraception.

**Funding:** This study was funded by a UK Medical Research Council (MRC) iCASE PhD studentship to Abigail Goodship (MR/W00710X/1).

## Introduction

Six years after the first identification of novel coronavirus SARS-CoV-2, coronavirus disease 2019 (COVID-19) continues to profoundly impact people’s lives. Whilst most SARS-CoV-2 infections resolve within 4 weeks, over 10% progress to a chronic multisystemic condition known as ‘long COVID’.^1^ Long COVID symptoms are wide-ranging, but commonly include ‘crashes’ (the sudden and severe worsening of symptoms following exertion), fatigue, brain fog and shortness of breath^2^ and can significantly reduce quality of life.^3^ By the end of 2023, there were an estimated 400 million global cases of long COVID, with true numbers likely higher. Recovery is slow, with only 7–10% fully recovered at two years post-infection.^4^

Approximately half of long COVID cases also meet the diagnostic criteria for myalgic encephalomyelitis (ME), also known as chronic fatigue syndrome (CFS).^5^ Whilst not all cases of ME/CFS have an identifiable trigger, many are preceded by a viral infection such as glandular fever,^6^ or Ross River virus.^7^ The prevalence of ME/CFS is estimated at 0.59% - 1.3%,^8–10^ and around a quarter of people living with ME/CFS are housebound.^11^ Despite the profound impact ME/CFS can have on quality of life, research has been underfunded.^12^ Given the historic neglect of ME/CFS, the recent emergence of long COVID, and the complexity of both, many questions about their pathophysiology remain unanswered.

Long COVID and ME/CFS both affect approximately twice as many females as males^13,14^ and higher oestrogen may be beneficial in females with acute COVID-19.^15,16^ Whilst the pathophysiology of long COVID and ME/CFS is still unclear, dysregulated immune responses may play a role.^17,18^ Sex hormones modulate sex differences in immune function^19^ and phases of the menstrual cycle correlate with symptoms of chronic, and particularly autoimmune, conditions often worsen at, or shortly before, menstruation^20^ (**appendix, p3**). Hormonal contraception alters cyclical fluctuations in ovarian hormones across the menstrual cycle and is associated with altered disease susceptibility and severity, ^20^ such as improved multiple sclerosis symptoms. ^21^

Cross-sectional surveys suggest that long COVID symptoms may also vary with the menstrual cycle,^2,22^ but these approaches can be affected by recruitment and recall bias. Many individuals track both menstrual cycles and disease symptoms using apps, entering data in real-time as part of their healthcare routine. Data collected in this way are less likely to be affected by recruitment bias than other methods of data collection.^23^

Visible is an app designed for people with chronic health conditions (such as long COVID and ME/CFS) to track their health, including symptoms and other factors such as menstruation.^24^ Here, we use mixed effects models to test the null hypotheses that 1) long COVID and ME/CFS symptoms do not differ between the menstrual period and other phases of the cycle, and 2) stabilising ovarian hormone levels with hormonal contraception is not associated with improved symptoms, rejecting the null when p<0.05. To determine the extent to which differences in symptoms over the menstrual cycle reflect fluctuations in wellbeing associated with normal menstrual cycles, we applied the same approaches to data from a general population of menstruating individuals using the Hertility menstrual tracker app^25^.

## Research in context

### Evidence before this study

Patients have reported premenstrual aggravation of ME/CFS for many years, yet research into both ME/CFS and long COVID and the menstrual cycle to date has been very limited, with existing studies limited to single self-reported questions on perceived symptom changes across the cycle. ^26^ Two small-scale studies have formally surveyed ME/CFS patients: 53% of 120 female participants in a 2019 study^27^ and 67% of the 42 cycling participants in a 2006 study^28^ reported worsening of their illness just before and during menstruation. For long COVID, a large, international patient-led survey from Davis et al. found that a third of menstruating participants (n =1792) reported relapses of their long COVID symptoms immediately before or during their period.^2^ In a smaller UK survey, 62% of female participants (n = 460) reported worsened long COVID symptoms on the days before their period.^22^ To our knowledge, the effect of hormonal contraception on ME/CFS and long COVID has yet to be formally investigated. In acute COVID-19, oestrogen may have a protective effect. A Swedish cohort study of post-menopausal women infected with SARS-CoV-2 found that higher oestrogen was associated with a lower risk of death.^15^ Furthermore, a population-based matched cohort study of UK women found that users of the combined hormonal contraceptive pill (n=295,689) had lower rates of COVID-19 hospital attendance.^16^

### Added value of this study

Previous research on menstrual cycle-related symptom changes in ME/CFS and long COVID has relied on retrospective surveys, capturing participants’ perceptions at a singular moment in time, and has also focused only on changes in the perimenstrual period. This study is the first to use daily, prospective, app-based tracking to quantify changes to long COVID and ME/CFS symptoms across the cycle, and to compare to symptom changes and to compare to symptom changes across the menstrual cycle in the general population. Symptoms were worse during the menstrual phase and less severe during the early luteal phase. This study is also the first to investigate the effect of hormonal contraception on symptoms of long COVID and ME/CFS. Use of combined hormonal contraception was associated with reduced risk of crashes and amelioration of other symptoms.

### Implications of all the available evidence

The finding that long COVID and ME/CFS symptoms follow a cyclical pattern over the menstrual cycle could empower people living with these conditions to schedule less strenuous activities during phases of the cycle when they are likely to be more vulnerable to crashes. The finding that combined hormonal contraceptives are associated with reduced symptoms could also inform the use of contraception in people living with long COVID and ME/CFS, although it should be noted that differences in health status or behaviours between hormonal contraceptive users and non-users may influence the observed associations with symptom severity. Our findings could suggest a role for sex hormones in disease modulation, although we do also observe cyclical reporting of some symptoms in the population comparison group.

## Methods

### Data source and collection

A retrospective analysis of symptom severity and menstrual cycle phase was conducted on data prospectively collected from users of the Visible app (the disease cohort). ^24^ On registration, app users enter their birth year, gender and whether they consider themselves to have long COVID, ME/CFS or another condition. Users who consented to participate in the study answered survey questions on their use of hormonal contraception, pregnancy status, breastfeeding status, number of pregnancies, and number of births. Each day, users self-report their commonly experienced symptoms on a 4-point Likert scale [None, Mild, Moderate, Severe] and whether they are currently experiencing a crash **(appendix, p31)**. The following definition was provided: “Crashes usually occur as a part of post-exertional symptom exacerbation and affect your ability to carry out your usual activities. They normally last a few days.’’ Reports entered between 7^th^ September 2022 to 6^th^ March 2024 were collected. Prospectively collected data from users of the Hertility menstrual cycle tracking app was used to create a comparison group of menstruating individuals in the general population (**appendix, p31-32**). *Data cleaning and menstrual cycle phase coding*

All data cleaning and subsequent analysis was conducted using R version 4.5.1 (2025-06-13) in RStudio. ^29^ For both disease and population cohorts, individuals outside the age range 18-45 were excluded because of the increased likelihood that they will have irregular cycles, confounding assignment of cycle phases^30^. Menstrual cycle phases were assigned based on the International Federation of Gynaecology and Obstetrics (FIGO) definition of the normal menstrual cycle^31^ (**appendix, p32**). For the disease cohort, users who did not complete the survey questions, or who were breastfeeding and/or pregnant, were excluded. Any users who did not have either long COVID or ME/CFS, did not track their periods, or did not ever report bleeding, were excluded. For the disease cohort, exclusion of users and cycles is summarised in **Figure 1B**; assignment of days to menstrual cycle phases in **Figure 1C**. Demographics of users at each filtering stage of the disease cohort cleaning are shown in **appendix (p3)**. Once menstrual cycle phases had been assigned to each day, symptom reports associated with that day were integrated into the dataset. No symptom tracking data was imputed. Symptoms reported retrospectively (>2 days after date) were removed to reduce recall bias. The exclusion of users and cycles for the population cohort is summarised in the **appendix (p4,32**).

**Figure 1:**
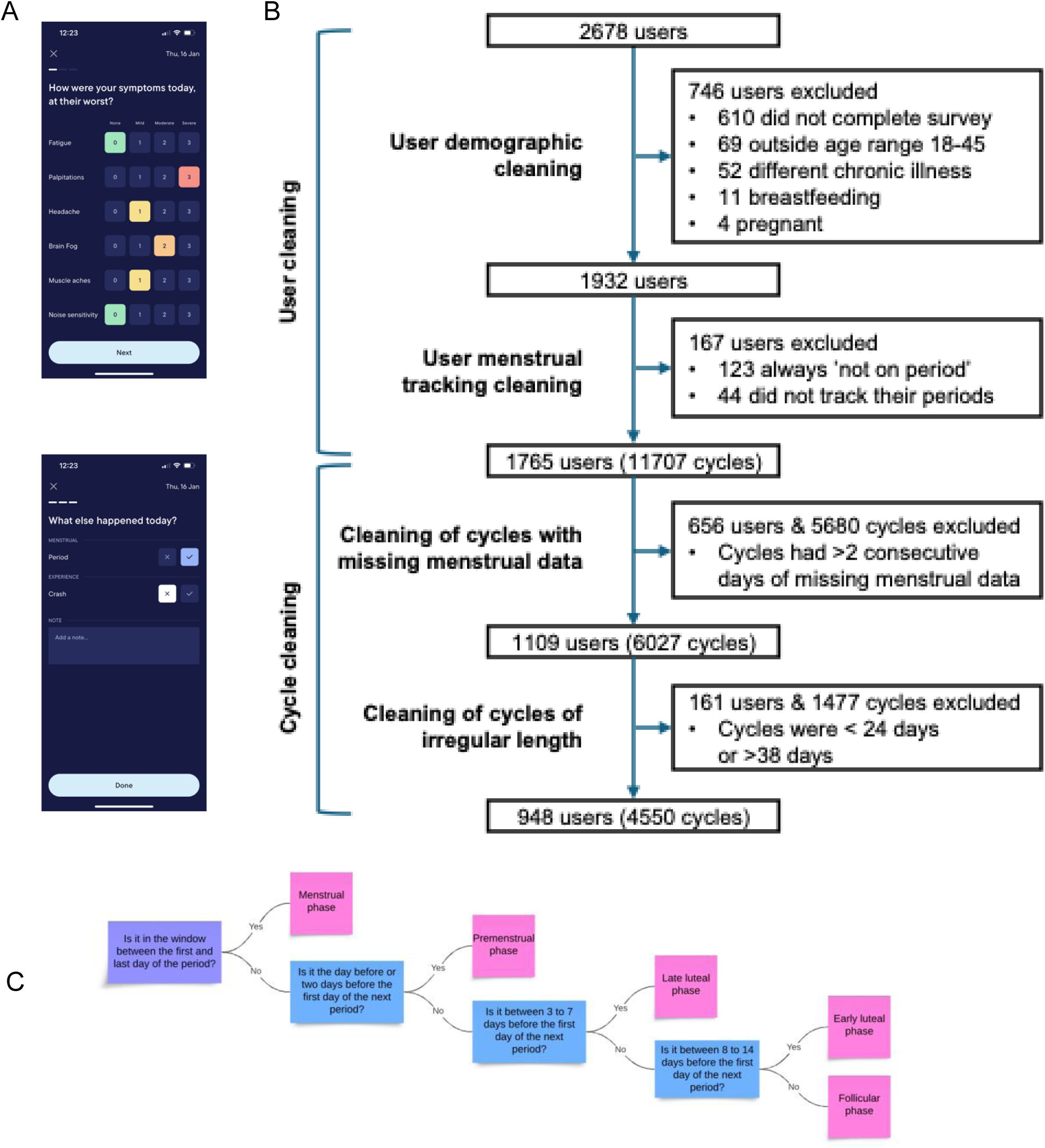
Overview of the study. (A) Screenshots from the Visible app illustrate how users track individual disease symptoms, menstruation and crashes. Data were collected from 7^th^ September 2022 to 6^th^ March 2024. (B) Flow diagram to illustrate the data cleaning process of users and menstrual cycles (C) The decision tree for assigning menstrual cycle phase to each day. Menstrual cycle phases were assigned based on the International Federation of Gynaecology and Obstetrics (FIGO) definition of the normal menstrual cycle^31^ (**appendix p32**).

### Descriptive statistics and statistical testing for disease cohort

Characteristics of the disease cohort are summarised in **Table 1** and descriptive plots are shown in **Figure 2**. Percentage of days classified as a crash was calculated out of the total days with data and displayed as a scatter plot with mean +/- standard error of the mean (SEM) for menstrual cycle phase, disease type and contraception type. Kruskal-Wallis tests were used to assess associations between mean crash rate and both disease type and contraception type. The Friedman test was used to test the significance between mean crash rate and menstrual phase.

**Figure 2:**
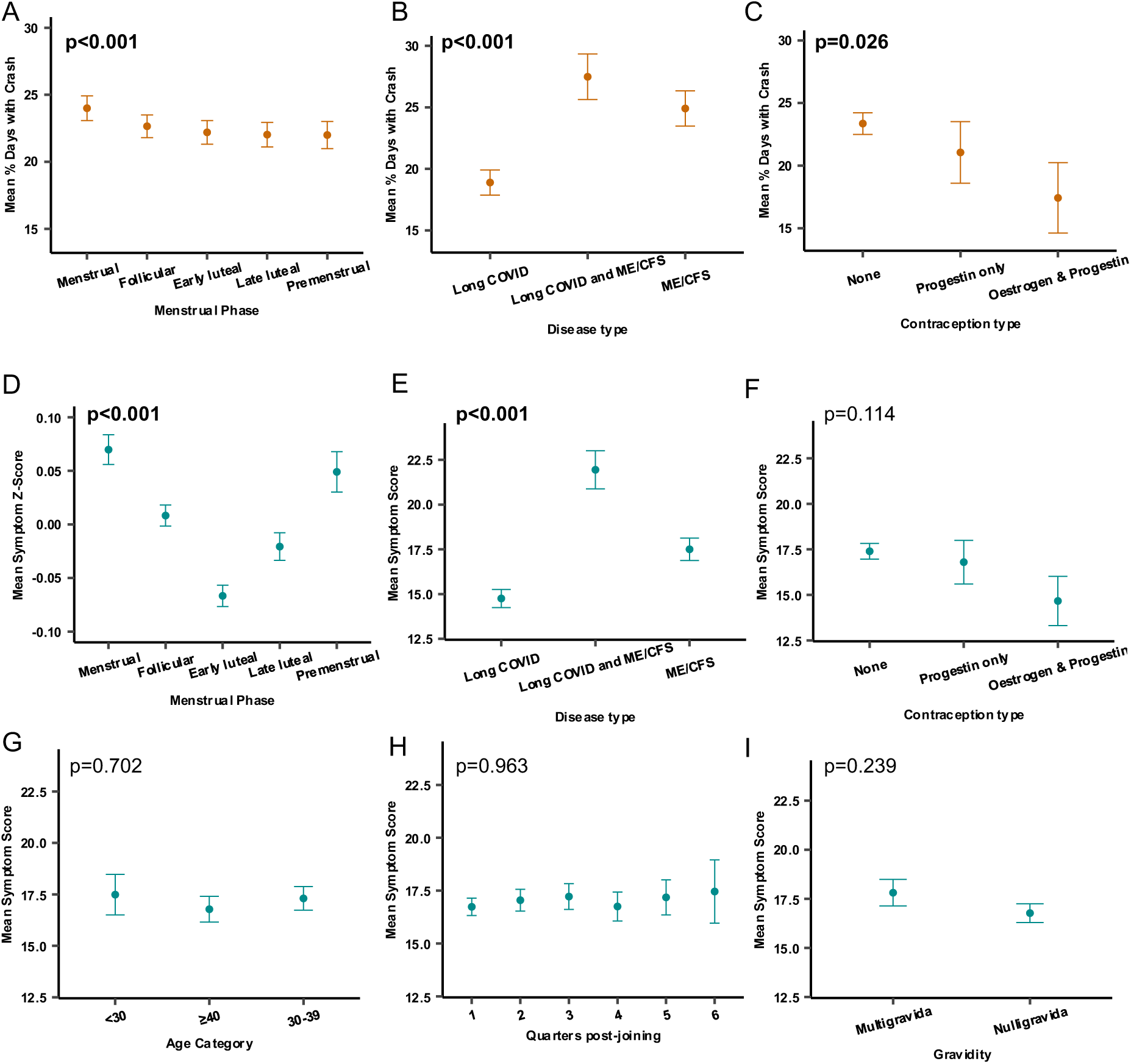
**The effects of menstrual phase, disease type, contraception type, time, gravidity and age on mean symptom severity** (A) Scatter plot showing mean percentage of days classified as a crash, displayed as mean +/- SEM, by menstrual phase. (B) Scatter plot showing mean percentage of days classified as a crash, displayed as mean +/- SEM, by disease type. (C) Scatter plot showing mean percentage of days classified as a crash, displayed as mean +/- SEM, by contraception type. (D-I) Mean symptom score was calculated by first summing the scores of each tracked symptom (all on a scale of 0-3) and then taking the mean across all days for each individual. Mean symptom Z-score was calculated by standardising each individual’s daily overall symptom score to their own mean and standard deviation (Z = (score − personal mean)/personal SD) and then averaging these Z-scores within each menstrual phase. Scatter plots show mean symptom Z-score by menstrual phase (D) and mean symptom score by disease type (E), contraception type (F), age category (G), quarters post-joining (H), and gravidity (I). Mean symptom score is displayed as mean +/- SEM. The Friedman test was used to test the significance between percentage crashes/mean symptom score and menstrual phase (A, D), due to the repeated measures nature of the data. Unpaired Kruskal-Wallis tests were used to test significance for disease type (B, E), contraception type (C, F), age category (G), quarters post-joining (H). The Mann-Whitney U-Test was used to test the significance between mean symptom score and gravidity (I), due to the binary nature of the variable. All p-values were extracted together and adjusted for multiple comparisons using the Bonferroni method, p<0.05 are highlighted in bold.

**Table 1.** Demographics of disease cohort.

| Variable | Subgroups | Count (n) | Percentage (%) |
| --- | --- | --- | --- |
| Disease type | Long COVID | 431 | 45.5 |
|  | ME/CFS | 329 | 34.7 |
|  | Long COVID and ME/CFS | 188 | 19.8 |
| Age category | <30 years old | 179 | 18.9 |
|  | 30-39 years old | 400 | 42.2 |
|  | ≥40 years old | 369 | 38.9 |
| Gender | Woman | 878 | 92.6 |
|  | Non-binary | 61 | 6.4 |
|  | Prefer not to say | 6 | 0.6 |
|  | Prefer to self-describe | 3 | 0.3 |
| Contraception type | None | 786 | 82.9 |
|  | Combined pill | 66 | 7.0 |
|  | Intrauterine system | 52 | 5.5 |
|  | Progestogen-only pill | 22 | 2.3 |
|  | Contraceptive implant | 18 | 1.9 |
|  | Patch | 4 | 0.4 |
| Number of times been pregnant | 0 | 617 | 65.1 |
|  | 1 | 87 | 9.2 |
|  | 2 | 132 | 13.9 |
|  | 3 | 112 | 11.8 |
| Number of times given birth | 0 | 687 | 72.5 |
|  | 1 | 71 | 7.5 |
|  | 2 | 135 | 14.2 |
|  | 3 | 55 | 5.8 |

All variables are displayed as count (n) and percentage (%) of the total users (N = 948)

For each Visible user, for each date, the scores of each tracked symptom were summed to give an overall symptom score and provide a comprehensive measure of a user’s overall health on a given day (36 symptoms, all on a scale of 0-3, theoretical range 0-108), reflecting the cumulative burden of symptoms. Incorporating the total number of symptoms experienced in addition to severity of symptoms is important, as for many individuals with long COVID, navigating the volume of symptoms feels “overwhelming and unmanageable”, although we acknowledge the potential for inflation of Type 1 error when aggregating scores for symptoms that may not be independent (Figures S12;S13).^32^

Mean overall symptom score was calculated for each individual by summing daily symptom scores and averaging across all days. Mean symptom Z-score was calculated by standardising each individual’s daily overall symptom score to their own mean and standard deviation (Z = (score − personal mean) / personal SD) and then averaging these standardised scores within each menstrual phase. These per-person phase means were used in the Friedman test to compare symptom scores across phases, ensuring that each participant contributed equally regardless of the number of cycles recorded. Kruskal-Wallis tests were used to assess the association between mean symptom score and disease type, contraception type, age category and quarters post-joining, defined as the number of three-month intervals since the participant registered with the app, used to account for potential changes in disease severity over time. Gravidity was assessed as a binary variable (nulligravida or not-nulligravida), and the Mann-Whitney U-Test was used to test the association between gravidity and mean symptom score. P values from all tests described above were extracted together and adjusted for multiple comparisons using the Bonferroni method. For R package details see **appendix (p33).**

### Mixed effects modelling for crashes, overall symptom score and individual symptoms

Generalised linear mixed-effects models assessed the impact of relevant variables on crash occurrence and overall symptom scores (disease cohort) and on individual symptoms (disease and population cohorts). Crash occurrence is a binary outcome, so a logistic regression (binomial family with a logit link) mixed-effects model was used. Fixed effects included menstrual phase, age category, disease type, contraception type, nulligravidity, and quarters since joining. ‘Individual’ was considered a random effect, which is recommended in menstrual cycle research to estimate between-person differences in within-person changes across the cycle.^30,33,34^ Model coefficients were exponentiated and expressed as odds ratios (ORs), which quantify the relative odds of a crash occurring for each predictor compared with the reference category, while accounting for covariates. An OR greater than 1 indicates higher odds of crash occurrence, whereas an OR less than 1 indicates lower odds, relative to the reference group.

To evaluate the effect of the same variables on overall symptom score, a negative binomial regression mixed-effects model was used. This approach was chosen because overall symptom score is a non-negative integer variable and exhibited overdispersion (variance greater than the mean), a common feature of count data. Results are reported as incidence rate ratios (IRRs), which describe how the expected symptom score changes relative to the reference group. An IRR greater than 1 indicates higher symptom scores, while an IRR less than 1 indicates lower symptom scores compared with the reference.

Mixed effects models were used to analyse fatigue, brain fog and headache in both cohorts. For the Visible disease cohort, symptoms were recorded on a 0–3 ordinal scale, and we therefore fitted cumulative link mixed-effects models (CLMMs) using the *clmm* function from the ordinal package.^35^ The models employed a logit link function and maximum likelihood estimation with Laplace approximation to account for random effects. The default optimiser (nlminb) was used, the maximum number of iterations set to 40,000, and the maximum absolute gradient of the inner optimisation to 1×10^-4^. For the population cohort, symptoms were recorded as binary (present/absent). Accordingly, we fitted mixed-effects logistic regression models using the glmer function from the lme4 package. Both models included ‘individual’ as a random effect and fixed effects of menstrual phase, age category, contraception type and gravidity. Model coefficients from both modelling frameworks were exponentiated and interpreted as ORs with 95% confidence intervals (CIs). Details of all models can be found in **appendix (p34-35).**

### Ethics

Ethical approval was obtained from the Research Governance and Integrity Team at Imperial College London, study numbers 6277771 and 8011320.

## Results

At study start date, 3297 Visible users were aged between 18 and 45 and had enabled the option to track their periods. 2678 Visible users consented for their anonymised data to be used in the study period (7^th^ September 2022 – 6^th^ March 2024), of whom 948 remained after data cleaning (**Figure 1B**, **Table 1**). Just under half the disease cohort identified as having long COVID only (45**·**5%), around a third as having ME/CFS only (34**·**7%), and a fifth (19**·**8%) as having both long COVID and ME/CFS. Almost all users identified as women (92**·**6%), with a smaller proportion identifying as non-binary (6.4%), six preferring not to say (0.6%), and three self-describing (0.3%). The majority of users were not on hormonal contraception (82**·**9%), and the remainder were split fairly evenly between oestrogen and progestin combined hormonal contraception (combined pill (7**·**0%) or patch (0**·**4%), hereafter called “combined hormonal contraception”) and progestin-only contraceptives (intrauterine system (5**·**5%) or progestogen-only pill (2**·**3%) or contraceptive implant (1**·**9%)). Around two-thirds of users had never been pregnant (65**·**1%) or given birth (72**·**5%). Median user age was 37 (IQR = 10, distribution in **appendix (p5)**). No significant differences were found between the characteristics of study participants by disease type **appendix (p6)**, but users on combined contraception were younger than users not on hormonal contraception or on progestin-only contraception (**appendix p6-7**).

Approximately 85% of people living with long COVID experience episodic symptoms that rapidly fluctuate from periods of stability to severe exacerbations, resulting in significant functional declines.^2^ These periods of severe worsening symptoms are often called “crashes” by those living with chronic illness,^36^ who may choose to avoid scheduling strenuous activities at times they are more vulnerable to crash. The mean percentage of days with crash was highest in the menstrual phase and decreased over the cycle, plateauing between the early luteal and premenstrual phases (**Figure 2A**). Adjusting for contraception type, disease type, age category, gravidity and time since joining the app using a mixed-effects logistic regression model, crashes remained less frequent in every phase compared to menstruation (**Table 2**). The late luteal phase showed the greatest reduction in crashes (OR=0**·**888, p<0·001). The follicular, early luteal and premenstrual phase also showed significant reductions (OR=0**·**947, p=0**·**037 and OR=0**·**900, p<0·001 and OR=0**·**919, p=0**·**033, respectively).

**Table 2.** The fitting result of binomial regression model (crashes)

| Fixed effects | Odds ratio<br>(OR) | 95% CI<br>(lower) | 95% CI<br>(upper) | P value |
| --- | --- | --- | --- | --- |
| **Menstrual phase** |  |  |  |  |
| Menses | - | - | - | - |
| Follicular | 0.947 | 0.900 | 0.997 | **0.037\*** |
| Early luteal | 0.900 | 0.853 | 0.949 | **P < 0.001\*** |
| Late luteal | 0.888 | 0.838 | 0.941 | **P < 0.001\*** |
| Premenstrual | 0.919 | 0.851 | 0.993 | **0.033\*** |
| **Hormonal contraception type** |  |  |  |  |
| None | - | - | - | - |
| Oestrogen & progestin | 0.548 | 0.350 | 0.856 | **0.008\*** |
| Progestin only | 0.864 | 0.593 | 1.260 | 0.446 |
| **Disease type** |  |  |  |  |
| Long COVID only | - | - | - | - |
| ME/CFS only | 1.468 | 1.141 | 1.889 | **0.003\*** |
| Long COVID and ME/CFS | 1.848 | 1.377 | 2.480 | **P < 0.001\*** |
| **Age category** |  |  |  |  |
| < 30 | - | - | - | - |
| 30-39 | 0.752 | 0.551 | 1.027 | 0.073 |
| >= 40 | 0.799 | 0.566 | 1.130 | 0.204 |
| **Gravidity** |  |  |  |  |
| Not-nulligravida | - | - | - | - |
| Nulligravida | 1.000 | 0.770 | 1.30 | 0.998 |
| **Quarters post-joining** |  |  |  |  |
| 1 | - | - | - | - |
| 2 | 0.943 | 0.894 | 0.994 | **0.030\*** |
| 3 | 0.937 | 0.881 | 0.997 | **0.040\*** |
| 4 | 1.097 | 1.023 | 1.176 | **0.009\*** |
| 5 | 0.979 | 0.903 | 1.061 | 0.597 |
| 6 | 1.090 | 0.851 | 1.396 | 0.495 |

\* P value <0.05
Random effect: Individual (intercept) Variance: 2.566 Standard deviation: 1.602

Crashes were significantly less frequent in users on combined hormonal contraception (OR=0**·**548, p=0·008), compared to users not on hormonal contraception (**Table 2**, **Figure 2B**). Users on progestin-only contraception also had fewer crashes than users not on hormonal contraception but this was not significantly different (OR=0**·**864, p=0**·**446). In comparison to those with long COVID, users with ME/CFS and with both long COVID and ME/CFS had significantly higher odds of crash (OR=1**·**468, p=0**·**003 and OR=1**·**848, P < 0·001) (**Table 2**, **Figure 2C**). Age and gravidity were not significantly associated with odds of crash.

To determine how overall disease severity changed over the cycle and with demographic characteristics, we examined the daily sum of all symptom ratings, the ‘symptom score’, with higher scores indicating higher severity. For each individual, we calculated the mean symptom score across all relevant days. The mean symptom z-score was lowest in the early luteal phase (mean=-0**·**067, SEM=0**·**01), significantly higher in the follicular (mean=0**·**008, SEM=0**·**01, p<0**·**001) and late luteal phases (mean=-0**·**021, SEM=0**·**013, p=0**·**003) and still higher in the menstrual (mean=0**·**067, SEM=0**·**014, p<0**·**001) and premenstrual phases (mean=0**·**049, SEM=0**·**019, p<0**·**001) (**Figure 2D**, see **appendix p8** for non-normalised graph). Mean symptom scores were highest in users with both long COVID and ME/CFS and were lowest in users with long COVID alone (**Figure 2E**). Mean symptom scores from users with different contraception types, time since joining, gravidity and age categories did not significantly differ in this analysis (**Figure 2F-I**).

Symptom score was positively skewed and over-dispersed (**appendix, p8**), so we used negative binomial multivariable mixed-effects regression and incorporated random intercepts for each user. The association between menstrual cycle phase and symptom score remained significant adjusting for contraception type, disease type, age category, gravidity and time since joining (**Table 3**). Symptom scores were significantly higher during menstruation than in all other cycle phases. Compared to the menstrual phase, the early luteal phase showed the greatest reduction in symptom scores (incidence rate ratio (IRR)=0·963, p<0·001); the follicular and late luteal phases also showed significant reductions (IRR=0·985, p<0·001 and IRR=0·980, p<0·001, respectively); and the reduction observed in the premenstrual phase was small, with only borderline significance (IRR=0**·**992, p=0**·**042). In the regression model, symptom scores were significantly lower in users of combined hormonal contraception (IRR=0**·**872, p=0**·**041), compared to users not on hormonal contraception. Users on progestin-only contraception also had lower symptom scores compared to users not on hormonal contraception, but the difference was not significantly different (IRR=0**·**985, p=0**·**849). Compared to those with long COVID, users with ME/CFS had significantly higher overall symptom scores (IRR=1**·**211, p<0·001), and users with both long COVID and ME/CFS had even higher overall symptom scores (IRR=1**·**525, p<0·001). Compared to reports submitted in the first three months since joining the app, symptom scores demonstrate a small but significant increase over time, with IRR = 1**·**057, p<0·001 by fifth quarter since joining. This was associated with an increase in the number of symptoms tracked over time **(appendix, p9).** Age and gravidity did not show a significant association with symptom scores.

**Table 3.** The fitting result of negative binomial regression model (overall symptom score)

| Fixed effects | Mean symptom score | Incidence rate ratio | 95% CI (lower) | 95% CI (upper) | P value |
| --- | --- | --- | --- | --- | --- |
| **Menstrual phase** |  |  |  |  |  |
| Menses | 17 | - | - | - | - |
| Follicular | 16.3 | 0.985 | 0.981 | 0.990 | **P < 0.001\*** |
| Early luteal | 16.2 | 0.963 | 0.958 | 0.968 | **P < 0.001\*** |
| Late luteal | 16.4 | 0.980 | 0.974 | 0.985 | **P < 0.001\*** |
| Premenstrual | 16.7 | 0.992 | 0.985 | 1.00 | **0.042\*** |
| **Hormonal contraception type** |  |  |  |  |  |
| None | 16.6 | - | - | - | - |
| Oestrogen & progestin | 16.0 | 0.827 | 0.690 | 0.992 | **0.041\*** |
| Progestin only | 14.3 | 0.985 | 0.839 | 1.155 | 0.849 |
| **Disease type** |  |  |  |  |  |
| Long COVID only | 14.2 | - | - | - | - |
| ME/CFS only | 16.3 | 1.211 | 1.089 | 1.347 | **P < 0.001\*** |
| Long COVID and ME/CFS | 21.7 | 1.525 | 1.343 | 1.731 | **P < 0.001\*** |
| **Age category** |  |  |  |  |  |
| < 30 | 18.5 | - | - | - | - |
| 30-39 | 15.8 | 0.982 | 0.860 | 1.122 | 0.790 |
| >= 40 | 16.2 | 0.913 | 0.789 | 1.057 | 0.223 |
| **Gravidity** |  |  |  |  |  |
| Not-nulligravida | 16.7 | - | - | - | - |
| Nulligravida | 15.9 | 0.911 | 0.815 | 1.018 | 0.099 |
| **Quarters post-joining** |  |  |  |  |  |
| 1 | 16.2 | - | - | - | - |
| 2 | 16.6 | 1.008 | 1.003 | 1.013 | **0.002\*** |
| 3 | 16.7 | 1.010 | 1.004 | 1.016 | **P < 0.001\*** |
| 4 | 15.9 | 1.039 | 1.032 | 1.045 | **P < 0.001\*** |
| 5 | 17.3 | 1.057 | 1.049 | 1.065 | **P < 0.001\*** |
| 6 | 15 | 1.049 | 1.025 | 1.074 | **P < 0.001\*** |

\* P value <0.05
Random effect: Individual (intercept) Variance: 0.542 Standard deviation: 0.736

The most tracked symptoms in the disease cohort were fatigue (99·5% of users), brain fog (88·3%) and headaches (85·1%) (**appendix p9**). These symptoms were frequently co-reported with each other and with muscle aches, dizziness, nausea, muscle weakness, joint pain, shortness of breath, palpitations, light sensitivity, noise sensitivity and memory issues **(appendix, p10-11)**. While crashes are specific to long COVID and ME/CFS, fatigue, headaches and muscle aches also occur in individuals without these conditions. To determine the extent to which the observed cyclicity is specific to long COVID/ME/CFS or instead reflects normal variation in wellbeing across the menstrual cycle, we therefore used data from a menstrual cycle tracking app, Hertility, to create a comparison cohort of menstruating individuals in the general population.

Severity and presence of symptoms show a similar pattern over the cycle in the disease (**Figure 3A-C**) and population (**Figure 3D-F**) cohorts, respectively, although the proportion of individuals reporting fatigue, brain fog and headaches is higher in the disease cohort. We used ordinal regression models to examine the association between menstrual cycle phase and the occurrence of fatigue, brain fog, and headache in both cohorts. In the disease cohort, odds of fatigue were significantly lower in the follicular (OR=0·947, p=0·002), early luteal (OR=0·860, p<0·001), late luteal (OR=0·908, p<0·001), and premenstrual phases (OR=0·915, p<0·001), compared to the menstrual phase (**Table 4A**). Similar patterns were observed for brain fog (**Table 4B**) and headache (**Table 4C**), with the largest reductions in the early luteal phase (brain fog OR=0·860, headache OR=0·567, both p<0·001). The population comparison cohort showed reductions in fatigue, brain fog and headache across all non-menstrual phases (all p<0·001) (**Tables 4A-C**). Hormonal contraception, age, and gravidity were not significantly associated with individual symptoms, except for a reduction in headache among those over 40 (OR=0·622, p<0·001) and nulligravida (OR=0·797, p=0·035) in the disease cohort. Overall, 32/36 individual symptoms tracked by the disease cohort showed a significant association between symptom severity and menstrual cycle phase, with 22 being worse in the menstrual phase and better in the luteal phase **(appendix, p12,17-28)** For symptoms that were also tracked in the population cohort, 9/9 showed a significant association between symptom severity and menstrual cycle phase, with all except diarrhoea most frequently present during menses **(appendix, p13,29-30)**

**Figure 3:**
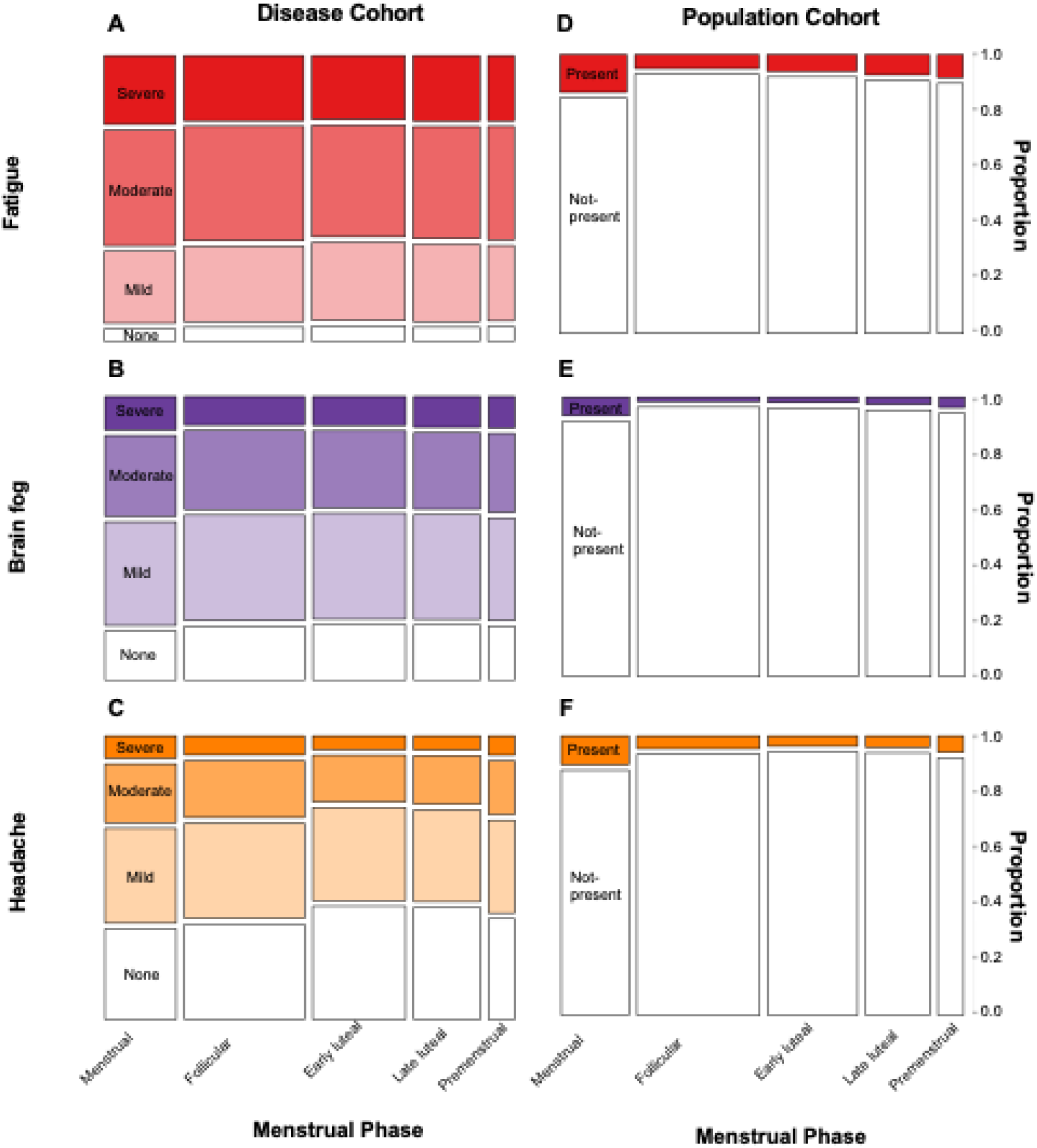
The impact of menstrual cycle phase on severity of the 3 most tracked symptoms in the disease cohort, compared to the population cohort. Individual symptom spineplots convey the frequencies of a two-way contingency table for the disease cohort (A-C) and the population comparison cohort (D-F). The widths of the bars correspond to the relative frequencies of reports within a menstrual cycle phase. The heights of the bars correspond to the conditional relative frequencies of the symptom severity rating in every menstrual cycle phase group. In the disease cohort, symptoms were reported on a 4-point Likert severity scale, in the population comparison cohort, symptoms were reported as present or not present.

**Table 4A.** The fitting result of fatigue regression models.

| Fixed effects | Disease<br>Odds ratio (95% CI) | P value | Population<br>Odds ratio (95% CI) | P value |
| --- | --- | --- | --- | --- |
| Menstrual phase |  |  |  |  |
| Menses | - | - | - | - |
| Follicular | 0.947 (0.916-0.980) | **0.002\*** | 0.289 (0.263-0.318) | **P < 0.001\*** |
| Early luteal | 0.860 (0.830-0.892) | **P < 0.001\*** | 0.334 (0.302-0.369) | **P < 0.001\*** |
| Late luteal | 0.908 (0.874-0.944) | **P < 0.001\*** | 0.424 (0.382-0.472) | **P < 0.001\*** |
| Premenstrual | 0.915 (0.869-0.963) | **P < 0.001\*** | 0.506 (0.440-0.583) | **P < 0.001\*** |
| Hormonal contraception type |  |  |  |  |
| None | - | - | - | - |
| Oestrogen & progestin | 0.821 (0.498-1.35) | 0.438 | 1.47 (0.830-2.60) | 0.187 |
| Progestin only | 1.08 (0.700-1.66) | 0.735 | 1.39 (0.886-2.18) | 0.152 |
| Age category |  |  |  |  |
| < 30 | - | - | - | - |
| 30-39 | 0.873 (0.621-1.23) | 0.432 | 1.19 (0.870-1.63) | 0.274 |
| >= 40 | 0.787 (0.546-1.14) | 0.202 | 1.53 (0.630-3.74) | 0.346 |
| Gravidity |  |  |  |  |
| Not-nulligravida | - | - | - | - |
| Nulligravida | 0.953 (0.715-1.27) | 0.744 | 1.43 (0.997-2.05) | 0.052 |

\* P value <0.05

**Table 4B.**
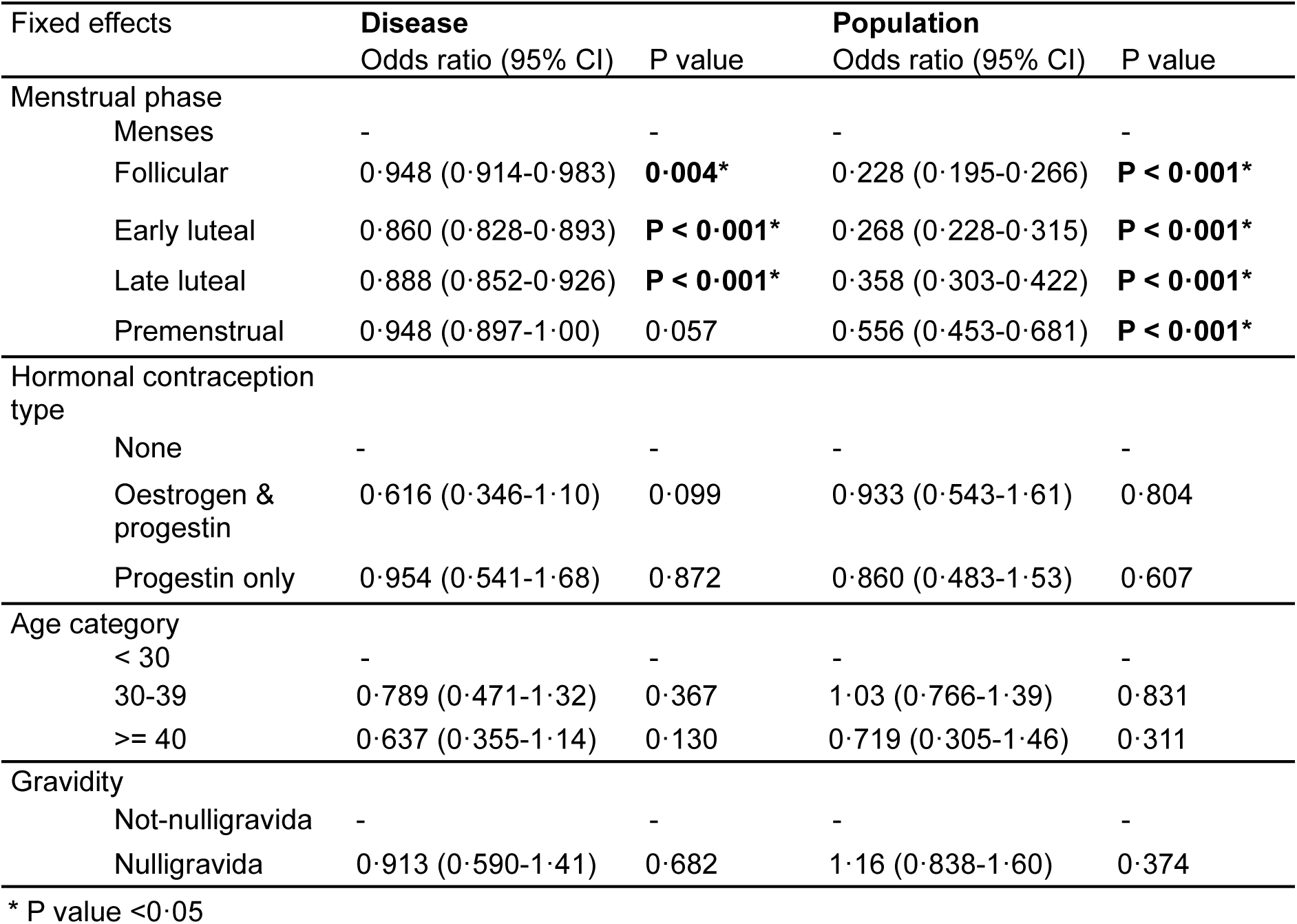
The fitting result of brain fog regression models.

| Fixed effects | Disease<br>Odds ratio (95% CI) | P value | Population<br>Odds ratio (95% CI) | P value |
| --- | --- | --- | --- | --- |
| Menstrual phase |  |  |  |  |
| Menses | - | - | - | - |
| Follicular | 0.948 (0.914-0.983) | **0.004\*** | 0.228 (0.195-0.266) | **P < 0.001\*** |
| Early luteal | 0.860 (0.828-0.893) | **P < 0.001\*** | 0.268 (0.228-0.315) | **P < 0.001\*** |
| Late luteal | 0.888 (0.852-0.926) | **P < 0.001\*** | 0.358 (0.303-0.422) | **P < 0.001\*** |
| Premenstrual | 0.948 (0.897-1.00) | 0.057 | 0.556 (0.453-0.681) | **P < 0.001\*** |
| Hormonal contraception type |  |  |  |  |
| None | - | - | - | - |
| Oestrogen & progestin | 0.616 (0.346-1.10) | 0.099 | 0.933 (0.543-1.61) | 0.804 |
| Progestin only | 0.954 (0.541-1.68) | 0.872 | 0.860 (0.483-1.53) | 0.607 |
| Age category |  |  |  |  |
| < 30 | - | - | - | - |
| 30-39 | 0.789 (0.471-1.32) | 0.367 | 1.03 (0.766-1.39) | 0.831 |
| >= 40 | 0.637 (0.355-1.14) | 0.130 | 0.719 (0.305-1.46) | 0.311 |
| Gravidity |  |  |  |  |
| Not-nulligravida | - | - | - | - |
| Nulligravida | 0.913 (0.590-1.41) | 0.682 | 1.16 (0.838-1.60) | 0.374 |

\* P value <0.05

**Table 4C.** The fitting result of headache regression models.

| Fixed effects | **Disease** |  | **Population** |  |
| --- | --- | --- | --- | --- |
|  | Odds ratio (95% CI) | P value | Odds ratio (95% CI) | P value |
| **Menstrual phase** |  |  |  |  |
| Menses | - | - | - | - |
| Follicular | 0.871 (0.866-0.876) | **P < 0.001\*** | 0.387 (0.349-0.428) | **P < 0.001\*** |
| Early luteal | 0.567 (0.564-0.571) | **P < 0.001\*** | 0.286 (0.252-0.322) | **P < 0.001\*** |
| Late luteal | 0.594 (0.590-0.597) | **P < 0.001\*** | 0.325 (0.286-0.369) | **P < 0.001\*** |
| Premenstrual | 0.797 (0.792-0.802) | **P < 0.001\*** | 0.510 (0.435-0.598) | **P < 0.001\*** |
| **Hormonal contraception type** |  |  |  |  |
| None | - | - | - | - |
| Oestrogen & progestin | 0.856 (0.533-1.37) | 0.520 | 1.70 (0.912-3.17) | 0.095 |
| Progestin only | 1.05 (0.695-1.59) | 0.816 | 1.12 (0.670-1.88) | 0.662 |
| **Age category** |  |  |  |  |
| < 30 | - | - | - | - |
| 30-39 | 0.830 (0.650-0.777) | 0.138 | 1.30 (0.954-1.77) | 0.097 |
| >= 40 | 0.622 (0.497-1.06) | **P < 0.001\*** | 1.49 (0.668-3.32) | 0.330 |
| **Gravidity** |  |  |  |  |
| Not-nulligravida | - | - | - | - |
| Nulligravida | 0.797 (0.646-0.984) | **0.035\*** | 1.22 (0.857-1.74) | 0.269 |

\* P value <0.05

## Discussion

This study is the first to use prospectively collected app data to describe changes to long COVID and ME/CFS symptoms across the menstrual cycle. In line with previous reports, crash incidence and symptom severity varied by disease^37^. The most tracked symptoms and clustering are broadly in line with Davis *et al.* 2021 who reported that fatigue and brain fog were the most frequent symptoms in long COVID patients, alongside post-exertional malaise/crashes^2^.

When disease type, age, reproductive history and contraception type were adjusted for, menstrual cycle phase had a small, but significant impact on crashes and symptom score, in line with previous reports^2,14^. Compared to menses, the likelihood of experiencing a crash on any given day in the late luteal phase was modestly lower (22.7% of menstrual days were crash days, compared with 20.7% of late luteal days). Differences in overall symptom scores across the menstrual cycle were statistically significant (IRRs ranged from 0.963 to 0.992, all p < 0.05) but the absolute changes were modest, ranging from 16.2 in the early luteal phase to 17.0 during menses. In symptom-specific analyses of the disease cohort, odds of fatigue, brain fog, and headache were lower in all non-menses phases, most notably the early luteal phase (fatigue OR=0·860, brain fog OR=0·860, headache OR=0·567, all p<0·001). The population comparison cohort showed even larger reductions across non-menstrual phases (all p<0·001), although symptoms affected a much smaller proportion of the population cohort (**Figure 3**). These findings indicate robust statistical effects, but the modest effect sizes mean that clinical impact should be interpreted with caution.

In the disease cohort, use of combined hormonal contraception was associated with significantly ameliorated symptoms and a reduced risk of crashes. Users on combined hormonal contraception had significantly fewer crashes (OR=0·548, p=0·008) and lower symptom scores (IRR=0·872, p=0·041) compared to non-users. For users not on hormonal contraception, 21.9% of all days are crash days, compared to 15.2% of days for users on combined contraception. This warrants further investigation as combined hormonal contraception is safe, cheap and widely available, making it a promising approach to ameliorate long COVID and ME/CFS symptoms.

The major strength of this study is its use of data prospectively collected from a large cohort of app users. Users enter data in real time as part of their personal healthcare routines, reducing recall and recruitment bias. The availability of a large volume of data enabled us to examine changes over the menstrual cycle in granular detail, since even the shortest phase (premenstrual) had sufficient user-days for statistical analysis. To our knowledge, this study also represents the first attempt to use prospectively-collected menstrual cycle tracking app data to define how wellbeing changes over the menstrual cycle in the general population.

However, several limitations should also be considered. Both cohorts represent a convenience sample, and app users may differ systematically from non-users. Individuals with limited digital access or literacy are unlikely to be app users, and those who choose to track their health might also have different characteristics or health behaviours compared to those who do not, potentially limiting generalisability.

In the disease cohort, disease status was self-reported, however limiting the cohort to only those with a formal diagnosis would have substantially restricted inclusion, given that only 1.0% of the 2.1 million individuals estimated to be living with long COVID in the UK in 2023 have a formal diagnosis,^38^ with higher diagnosis rates among white individuals and those in less deprived areas^39^. Use of self-reported status maximises inclusivity and reduces risk of systematic bias, but findings should be interpreted with this limitation in mind. An additional limitation is the absence of information on how long participants had been living with their chronic illness, as disease course and symptom manifestations can change over time.

Daily symptoms are also self-reported, and users might rate symptoms differently. To address this, we included ‘individual’ as a random effect in the model. However, symptom perception biases may still influence patterns of reporting, particularly in the context of the menstrual cycle. Individuals may be more attuned to bodily sensations at certain phases of the cycle, such as during menstruation or the premenstrual period, which could lead to heightened reporting of symptoms. Conversely, some individuals may under-report symptom exacerbations during the perimenstrual phase if they consider these to be associated with menstruation, rather than their chronic illness. Our finding that, in the general population, individuals are also more likely to report fatigue, brain fog and headache at the time of their period suggests that some of the cyclicity in symptom reporting in the disease cohort may reflect normal variation across the menstrual cycle. However, that frequency of crash, which is not reported by healthy individuals, also varies across the cycle suggests that at least some aspects of cyclical symptom variation we report are disease specific.

To minimise the cognitive burden of participation on our disease cohort, we designed the survey to be as short as possible, but this meant that we did not collect information on potential confounders, including body-mass index, comorbidities, medication use, socioeconomic status, ethnicity, geographic context and disease duration. Within-individual analysis means that the association between symptom severity and menstrual cycle phase is likely to be robust to this, but this could certainly affect the relationship between contraceptive use and symptoms, and for this analysis we cannot rule out reverse causality. For example, combined contraceptive users might be generally healthier or more proactive in managing their health. Furthermore, the survey questions were completed when the user first enrolled so any changes in contraception use during the study were not captured. Nor did we collect information on use of emergency contraception, testosterone or hormone replacement therapy.

Assigning menstrual cycle phases based on menstrual period dates is most accurate in individuals with regular cycles.^31^ However, this excludes individuals with absent or irregular cycles, which are reported by approximately a third of menstruating people living with long COVID,^2^ and could affect the representativeness of our findings. Even among regularly cycling individuals, ovarian ultrasound and blood or urine luteinising hormone measurements allow more accurate assignment of cycle phases, but data on these were not available, potentially introducing some inaccuracy in cycle phase assignment.

The results presented here support the idea that menstrual cycle phase influences long COVID and ME/CFS symptoms. The knowledge that they are more likely to crash during menstruation could allow people living with long COVID or ME/CFS to schedule strenuous activities in other phases of the menstrual cycle. Furthermore, the association between combined hormonal contraceptives, lower symptom scores and fewer crashes points towards a role for ovarian hormones in modulating symptoms and highlights the therapeutic potential of oestrogen-containing contraceptives or hormone replacement therapy, that could be tested in randomised studies. Future work should also investigate how these conditions intersect with other aspects of female reproductive health. For example, understanding how pregnancy influences long COVID symptoms could help guide family planning decisions. Connecting symptom patterns with ovarian hormone dynamics sets the stage for targeted interventions and precision care in long COVID and ME/CFS.

## Declarations

### Availability of data and materials

While all data used in this analysis were anonymised, the individual-level nature of the data used risks individuals being identified or being able to self-identify if it is released publicly. However, the data can be requested by bona fide researchers for specified scientific purposes by emailing for the disease cohort data and for the population cohort data.

### Code availability

https://github.com/AbiGoodship/effect_of_ovarian_hormones_on_long_COVID_and_ME_symptoms

### Authors’ Contributions

VM and CM conceptualised the study. RP and HL curated Visible data; HON and NG curated Hertility data. JH, AG and CM developed the methodology. AG formally analysed and visualised the data. CM and VM supervised the study. AG wrote the original manuscript draft. All authors were responsible for the review and editing of the manuscript. All authors discussed, edited, and approved the final version of the manuscript. All authors had final responsibility for the decision to submit the manuscript for publication.

## Data Availability

While all data used in this analysis were anonymised, the individual-level nature of the data used risks individuals being identified or being able to self-identify if it is released publicly. However, the data can be requested by bona fide researchers for specified scientific purposes by emailing for the disease cohort data and for the population cohort data.
Code is available at https://github.com/AbiGoodship/effect_of_ovarian_hormones_on_long_COVID_and_ME_symptoms

## References

1 Davis HE, McCorkell L, Vogel JM, Topol EJ. Long COVID: major findings, mechanisms and recommendations. Nat Rev Microbiol. 2023; 21. DOI:10.1038/s41579-022-00846-2.

2 Davis HE, Assaf GS, McCorkell L, et al. Characterizing long COVID in an international cohort: 7 months of symptoms and their impact. EClinicalMedicine 2021; 38. DOI:10.1016/j.eclinm.2021.101019.

3 Carlile O, Briggs A, Henderson AD, et al. Impact of long COVID on health-related quality-of-life: an OpenSAFELY population cohort study using patient-reported outcome measures (OpenPROMPT). The Lancet Regional Health - Europe 2024; 40: 100908.

4 Al-Aly Z, Davis H, McCorkell L, et al. Long COVID science, research and policy. Nature Medicine 2024 30:8 2024; 30: 2148–64.

5 Dehlia A, Guthridge MA. The persistence of myalgic encephalomyelitis/chronic fatigue syndrome (ME/CFS) after SARS-CoV-2 infection: A systematic review and meta-analysis. Journal of Infection 2024; 89: 106297.

6 Katz BZ, Shiraishi Y, Mears CJ, Binns HJ, Taylor R. Chronic Fatigue Syndrome After Infectious Mononucleosis in Adolescents. Pediatrics 2009; 124: 189–93.

7 Hickie I, Davenport T, Wakefield D, et al. Post-infective and chronic fatigue syndromes precipitated by viral and non-viral pathogens: prospective cohort study. BMJ 2006; 333: 575.

8 Valdez AR, Hancock EE, Adebayo S, et al. Estimating prevalence, demographics, and costs of ME/CFS using large scale medical claims data and machine learning. Front Pediatr 2019; 6: 433309.

9 Vahratian A, Lin J-MS, Bertolli J, Unger ER. Myalgic Encephalomyelitis/Chronic Fatigue Syndrome in Adults: United States, 2021-2022. 2023; published online Dec 13. DOI:10.15620/CDC:134504.

10 Samms GL, Ponting CP. Unequal access to diagnosis of myalgic encephalomyelitis in England. medRxiv 2024; : 2024.01.31.24302070.

11 Pendergrast T, Brown A, Sunnquist M, et al. Housebound versus nonhousebound patients with myalgic encephalomyelitis and chronic fatigue syndrome. Chronic Illn 2016; 12: 292–307.

12 Smith K. Women’s health research lacks funding — these charts show how. Nature 2023; 617. DOI:10.1038/d41586-023-01475-2.

13 Lim EJ, Ahn YC, Jang ES, Lee SW, Lee SH, Son CG. Systematic review and meta-analysis of the prevalence of chronic fatigue syndrome/myalgic encephalomyelitis (CFS/ME). J Transl Med 2020; 18. DOI:10.1186/S12967-020-02269-0.

14 Sigfrid L, Drake TM, Pauley E, et al. Long Covid in adults discharged from UK hospitals after Covid-19: A prospective, multicentre cohort study using the ISARIC WHO Clinical Characterisation Protocol. The Lancet Regional Health - Europe 2021; 8. DOI:10.1016/j.lanepe.2021.100186.

15 Sund M, Fonseca-Rodríguez O, Josefsson A, Welen K, Fors Connolly AM. Association between pharmaceutical modulation of oestrogen in postmenopausal women in Sweden and death due to COVID-19: A cohort study. BMJ Open 2022; 12. DOI:10.1136/bmjopen-2021-053032.

16 Costeira R, Lee KA, Murray B, et al. Estrogen and COVID-19 symptoms: Associations in women from the COVID Symptom Study. PLoS One 2021; 16. DOI:10.1371/journal.pone.0257051.

17 Sukocheva OA, Maksoud R, Beeraka NM, et al. Analysis of post COVID-19 condition and its overlap with myalgic encephalomyelitis/chronic fatigue syndrome. J Adv Res. 2022; 40. DOI:10.1016/j.jare.2021.11.013.

18 Altmann DM, Whettlock EM, Liu S, Arachchillage DJ, Boyton RJ. The immunology of long COVID. Nat Rev Immunol. 2023; 23. DOI:10.1038/s41577-023-00904-7.

19 Hoffmann JP, Liu JA, Seddu K, Klein SL. Sex hormone signaling and regulation of immune function. Immunity. 2023; 56. DOI:10.1016/j.immuni.2023.10.008.

20 Alvergne A, Högqvist Tabor V. Is Female Health Cyclical? Evolutionary Perspectives on Menstruation. Trends Ecol Evol. 2018; 33. DOI:10.1016/j.tree.2018.03.006.

21 Kempe P, Hammar M, Brynhildsen J. Symptoms of multiple sclerosis during use of combined hormonal contraception. European Journal of Obstetrics and Gynecology and Reproductive Biology 2015; 193. DOI:10.1016/j.ejogrb.2015.06.030.

22 Newson L, Lewis R, O’Hara M. Long Covid and menopause - the important role of hormones in Long Covid must be considered. Maturitas 2021; 152. DOI:10.1016/j.maturitas.2021.08.026.

23 Milne-Ives M, Van Velthoven MH, Meinert E. Mobile apps for real-world evidence in health care. Journal of the American Medical Informatics Association. 2020; 27. DOI:10.1093/jamia/ocaa036.

24 Visible - Activity tracking for Long Covid and ME/CFS. https://www.makevisible.com/ (accessed Aug 2, 2024).

25 At-Home Hormone & Fertility Tests For Women | Hertility Health. https://hertilityhealth.com/ (accessed Nov 27, 2025).

26 Pollack B, von Saltza E, McCorkell L, et al. Female reproductive health impacts of Long COVID and associated illnesses including ME/CFS, POTS, and connective tissue disorders: a literature review. Frontiers in Rehabilitation Sciences. 2023; 4. DOI:10.3389/fresc.2023.1122673.

27 Chu L, Valencia IJ, Garvert DW, Montoya JG. Onset patterns and course of myalgic encephalomyelitis/chronic fatigue syndrome. Front Pediatr 2019; 7: 427132.

28. Briggs N, Griffith L. Myalgic Encephalopathy/Chronic Fatigue Syndrome Longitudinal Outcomes. 2006.

29 Posit team. RStudio: Integrated Development Environment for R. 2024.

30 Schmalenberger KM, Tauseef HA, Barone JC, et al. How to study the menstrual cycle: Practical tools and recommendations. Psychoneuroendocrinology. 2021; 123. DOI:10.1016/j.psyneuen.2020.104895.

31 Munro MG, Critchley HOD, Fraser IS, et al. The two FIGO systems for normal and abnormal uterine bleeding symptoms and classification of causes of abnormal uterine bleeding in the reproductive years: 2018 revisions. International Journal of Gynecology and Obstetrics 2018; 143. DOI:10.1002/ijgo.12666.

32 Wurz A, Culos-Reed SN, Franklin K, DeMars J, Wrightson JG, Twomey R. ‘I feel like my body is broken’: exploring the experiences of people living with long COVID. Quality of Life Research 2022; 31. DOI:10.1007/s11136-022-03176-1.

33 Gehlert S, Song IH, Chang CH, Hartlage SA. The prevalence of premenstrual dysphoric disorder in a randomly selected group of urban and rural women. Psychol Med 2009; 39. DOI:10.1017/S003329170800322X.

34 Wei SM, Schiller CE, Schmidt PJ, Rubinow DR. The role of ovarian steroids in affective disorders. Curr Opin Behav Sci. 2018; 23. DOI:10.1016/j.cobeha.2018.04.013.

35 Christensen RHB. ordinal---Regression Models for Ordinal Data. 2023. https://CRAN.R-project.org/package=ordinal (accessed Jan 22, 2025).

36 O’Brien KK, Brown DA, McDuff K, et al. Conceptualising the episodic nature of disability among adults living with Long COVID: a qualitative study. BMJ Glob Health 2023; 8. DOI:10.1136/BMJGH-2022-011276.

37 Haider S, Janowski AJ, Lesnak JB, et al. A comparison of pain, fatigue, and function between post-COVID-19 condition, fibromyalgia, and chronic fatigue syndrome: a survey study. Pain 2023; 164: 385–401.

38 Prevalence of ongoing symptoms following coronavirus (COVID-19) infection in the UK - Office for National Statistics. https://www.ons.gov.uk/peoplepopulationandcommunity/healthandsocialcare/conditionsanddiseases/bulletins/prevalenceofongoingsymptomsfollowingcoronaviruscovid19infectionintheuk/5january2023 (accessed Sept 1, 2025).

39 Henderson AD, Butler-Cole BF, Tazare J, et al. Clinical coding of long COVID in primary care 2020–2023 in a cohort of 19 million adults: an OpenSAFELY analysis. EClinicalMedicine 2024; 72: 102638.

