## Supplementary information for "Digital health app data reveals an effect of ovarian hormones on long COVID and myalgic encephalomyelitis symptoms"

### Supplementary Figures and Tables

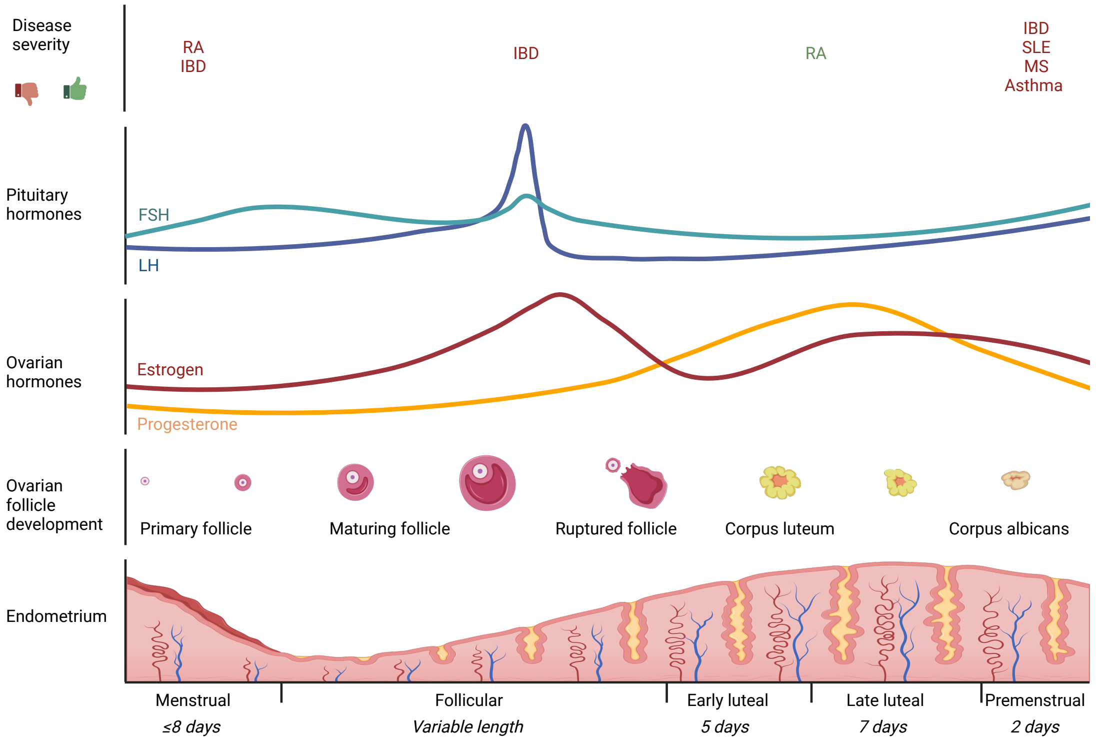

**Figure S1: Changes over the menstrual cycle**
A schematic representation of the menstrual cycle illustrates relative changes in pituitary and ovarian hormones, follicle development, and endometrial lining. Correlated changes in disease severity and infections are shown in the top panel, red indicates disease symptoms worsen/susceptibility to infection increases and green indicates disease symptoms improve/response to infection improves. Created in Biorender.com.[1]

| | | N=1932 | N=1765 | N=1109 | N=948 | P value |
|---|---|---|---|---|---|---|
| **Disease group (%)** | **Long COVID** | 40.17 | 41.08 | 44.45 | 45.46 | **0.026*** |
| | **ME/CFS** | 40.84 | 39.43 | 36.43 | 34.70 | |
| | **Both long COVID and ME/CFS** | 19.0 | 19.49 | 19.12 | 19.83 | |
| **Contraception type (%)** | **None** | 78.38 | 79.28 | 80.23 | 82.91 | 0.170 |
| | **Oestrogen & progestin** | 9.02 | 8.34 | 8.48 | 7.38 | |
| | **Progestin only** | 12.60 | 12.37 | 11.28 | 9.70 | |
| **Nulligravida (%)** | | 63.3 | 62.78 | 64.56 | 65.08 | 0.588 |
| **Age (mean)** | | 35.40 | 35.61 | 35.1 | 36.11 | **0.024*** |
| **Daily symptom score (mean)** | | 17.35 | 17.34 | 16.74 | 16.45 | **<0.001*** |
| **Percentage of days with crash (mean)** | | 32.1 | 32.1 | 22.9 | 22.7 | **<0.001*** |

**Table S1: Demographics of each filtering stage of the disease cohort cleaning**
1932 users filled out the survey, were within the age range of 18-45, had long COVID and/or ME/CFS, and were not breastfeeding and/or pregnant. The 1932 users were filtered down to 1765 users by removing any users who did not track their periods or report any bleeding days. The 1765 users were then filtered down to 1109 users by excluding cycles that had >2 consecutive days of missing menstrual data. The 1109 users were then filtered down to 948 users by excluding cycles that were abnormally short (<24 days) or long (>38 days). For each of these groups, the following demographic indicators were reported % in each disease group, % each contraception type, % nulligravida, mean age, mean daily symptom score, mean % of days with crash. Chi-squared tests were conducted to compare numbers in each disease group, contraception type and nulligravida. Kruskal-Wallis tests were conducted to compare age, daily symptom score and crash rate.

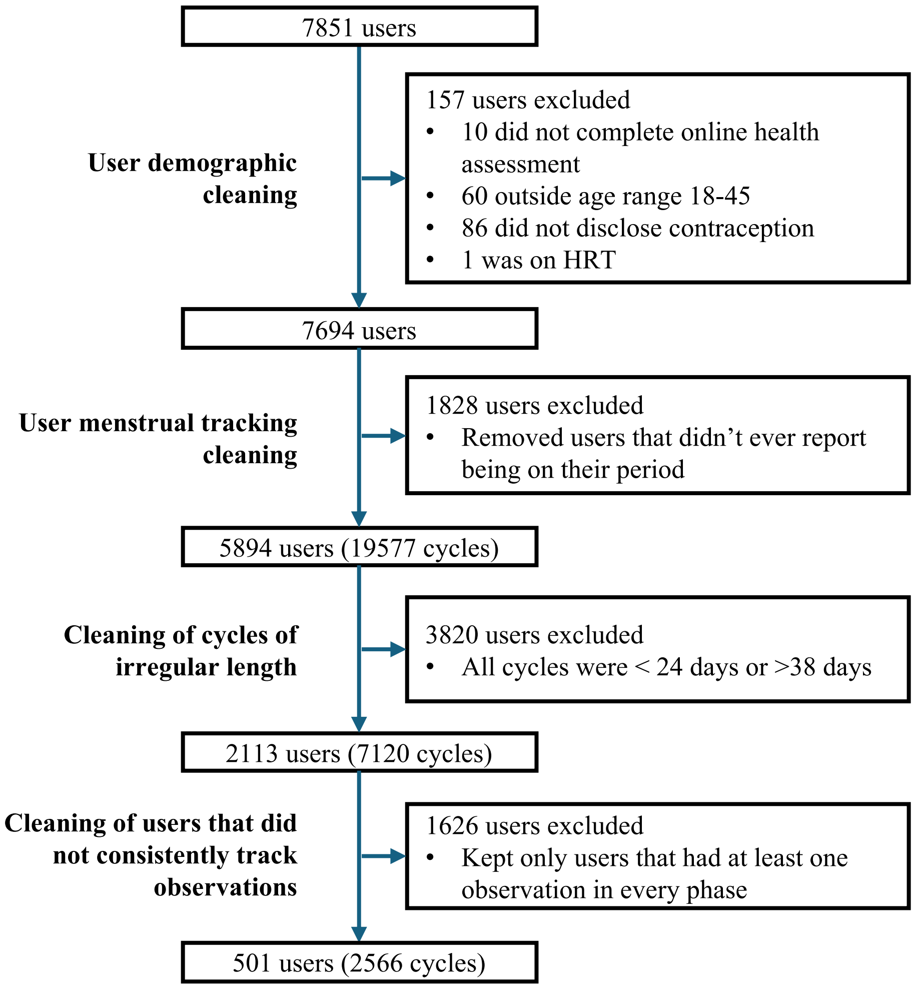

**Figure S2: Cleaning of the population cohort**
Flow diagram to illustrate the data cleaning process of users and menstrual cycles in the population cohort. Data obtained from the Hertility menstrual cycle tracking app (7th September 2022 -28th September 2025).

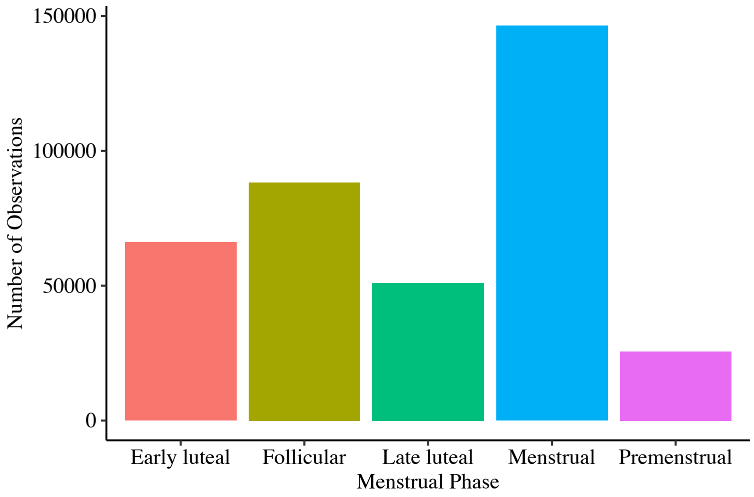

**Figure S3: Number of observations reported in each menstrual phase (before cleaning of users who did not consistently track observations) in the population cohort**
Bar plot showing the total number of symptom-tracking observations recorded across menstrual cycle phases after excluding flow-related entries (users=2113), but before cleaning of users that did not consistently track observations. Data represent the count of all non-flow symptoms logged within each phase, illustrating variation in participant engagement across the cycle (see **page 31** for full list of non-flow symptoms).

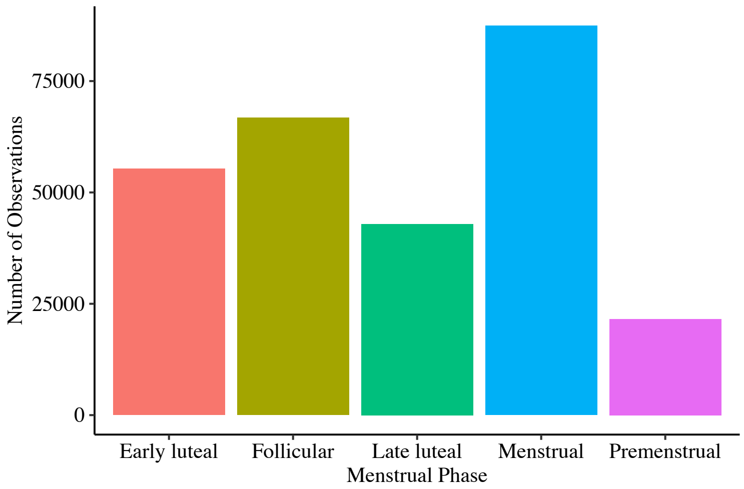

**Figure S4: Number of observations reported in each menstrual phase (after cleaning of users who did not consistently track observations) in the population cohort**
Bar plot showing the total number of symptom-tracking observations recorded across menstrual cycle phases after excluding flow-related entries and after excluding users who did not have at least one non-flow observation in each phase (users=501). Data represent the count of all non-flow symptoms logged within each phase, illustrating how variation in participant engagement across the cycle has decreased (see **page 31** for full list of non-flow symptoms).

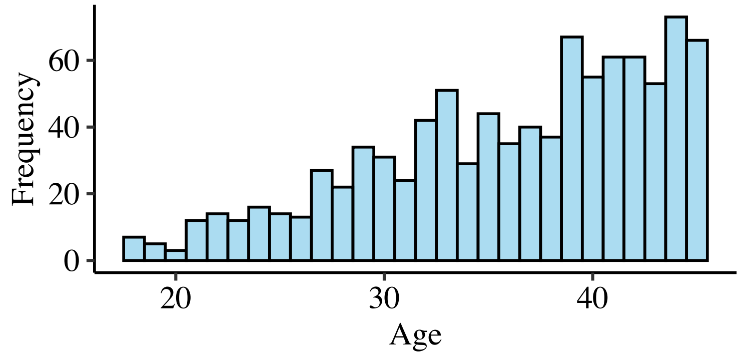

**Figure S5: Age distribution of the disease cohort**
Shown as a histogram, (N=948), bin width 1. Data obtained from the Visible app (7$^{th}$ September 2022 -6$^{th}$ March 2024).

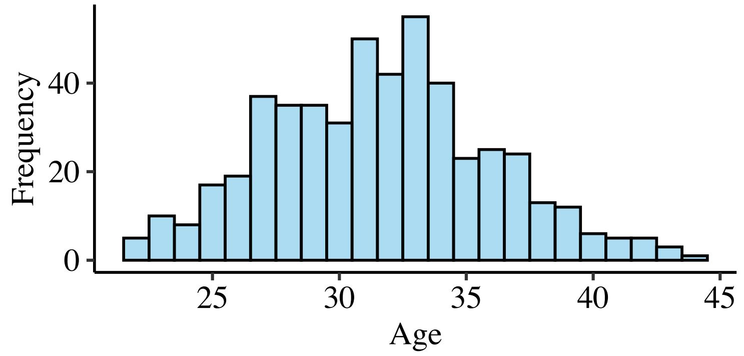

**Figure S6: Age distribution of the population cohort**
Shown as a histogram, (N=501), bin width 1. Data obtained from the Hertility menstrual cycle tracking app (7$^{th}$ September 2022 -28$^{th}$ September 2025).

**Table S2: Demographics of disease population, split by disease type**

| | Long COVID (n = 431) | ME/CFS (n = 329) | Long COVID & ME/CFS (n = 188) | P-value* |
|---|---|---|---|---|
| **Age (mean (IQR))** | 38 (10) | 37 (11) | 38(11) | 0.3316 |
| **Gender** | | | | |
| **Woman** | 409 (94·9) | 296 (90·0) | 173 (92·0) | 0.2238 |
| **Non-binary** | 17 (3·9) | 30 (9·1) | 14 (7·4) | |
| **Prefer not to say** | 4 (0·9) | 1 (0·3) | 1 (0·5) | |
| **Prefer to self-describe** | 1 (0·2) | 2 (0·6) | 0 (0.0) | |
| **Contraception type** | | | | |
| **None** | 354 (82·1) | 270 (82·1) | 162 (86·2) | 0.7358 |
| **Oestrogen & progestin** | 34 (7·9) | 26 (7·9) | 10 (5·3) | |
| **Progestin only** | 43 (10·0) | 33 (10·0) | 16 (8·5) | |
| **Number of times been pregnant** | | | | |
| **0** | 267 (61·9) | 226 (68·7) | 124 (66·0) | 0.5676 |
| **1** | 41 (9·5) | 29 (8·8) | 17 (9·0) | |
| **2** | 67 (15·5) | 42 (12·8) | 23 (12·2) | |
| **3** | 56 (13·0) | 32 (9·7) | 24 (12·8) | |
| **Number of times given birth** | | | | |
| **0** | 297 (68·9) | 253 (76·9) | 137 (72·9) | 0.09987 |
| **1** | 33 (7·7) | 22 (6·7) | 16 (8·5) | |
| **2** | 77 (17·9) | 37 (11·2) | 21 (11·2) | |
| **3** | 24 (5·6) | 17 (5·2) | 14 (7·4) | |

Excluding age, all parameters are displayed as n (%) of the total users (N=948).
Age is displayed as median number of years (interquartile range).
*Kruskal-Wallis rank sum test (age) or Pearson's $\chi^2$ test (all other parameters).

**Table S3: Demographics of disease population, split by contraception type**

| | No hormonal contraception (n = 786) | Progestin-only contraception (n = 92) | Combined contraception (n = 70) | P-value* |
|---|---|---|---|---|
| **Age (mean (IQR))** | 38 (10) | 38·5 (12) | 34 (10) | **0.00509** |
| **Gender** | | | | |
| **Woman** | 727 (92·5) | 86 (93·5) | 65 (92·9) | 0.9264 |
| **Non-binary** | 50 (6·4) | 6 (6·5) | 5 (7·1) | |
| **Prefer not to say** | 6 (0·8) | 0 (0·0) | 0 (0·0) | |
| **Prefer to self-describe** | 3 (0·4) | 0 (0·0) | 0 (0.0) | |
| **Disease type** | | | | |
| **Long COVID** | 354 (45·0) | 43 (46·7) | 34 (48·6) | 0.7358 |
| **ME/CFS** | 270 (34·4) | 33 (35·9) | 26 (37·1) | |
| **Both diseases** | 162 (20·6) | 16 (17·4) | 10 (14·3) | |
| **Number of times been pregnant** | | | | |
| **0** | 506 (64·4) | 55 (59·8) | 56 (80·0) | 0.1821 |
| **1** | 73 (9·3) | 10 (10·9) | 4 (5·7) | |
| **2** | 111 (14·1) | 14 (15·2) | 7 (10·0) | |
| **3** | 96 (12·2) | 13 (14·1) | 3 (4·3) | |
| **Number of times given birth** | | | | |
| **0** | 568 (72·3) | 61 (66·3) | 58 (82·9) | 0.3760 |
| **1** | 60 (7·6) | 7 (7·6) | 4 (5·7) | |
| **2** | 111 (14·1) | 18 (19·6) | 6 (8·6) | |
| **3** | 47 (6·0) | 6 (6·5) | 2 (2·9) | |

Excluding age, all parameters are displayed as n (%) of the total users (N=948).
Age is displayed as median number of years (interquartile range).
*Kruskal-Wallis rank sum test (age) or Pearson's $\chi^2$ test (all other parameters).

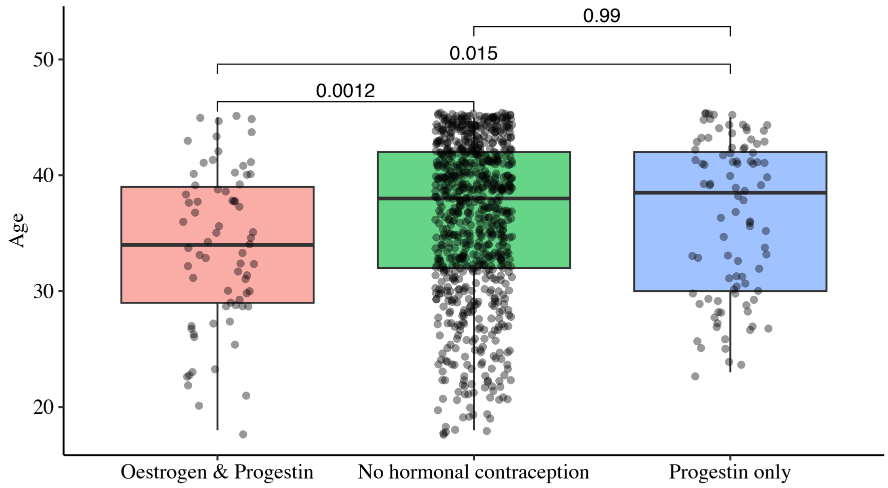

**Figure S7: Age distribution across contraception types, in the disease cohort**
Age shown as a boxplot, split by contraception type (users not on hormonal contraception (N=786), users on progestin-only contraception (N=92), users on combined oestrogen & progestin contraception (N=70)). Individual data points are overlaid using jittered scatter to show sample density within each group. Boxes represent the interquartile range (IQR), with the median indicated by a horizontal line; whiskers extend to 1.5× the IQR. Kruskal-Wallis tests were used to assess the association between age and disease type. Post-hoc pairwise comparisons were conducted with Dunn's tests, and p-values were adjusted using the Bonferroni method to control for Type 1 error.

**Table S4: Demographics of population cohort**

| Variable | Subgroups | Count (n) | Percentage (%) |
|---|---|---|---|
| **Age category** | <30 years old | 166 | 33.1 |
| | 30-39 years old | 315 | 62.9 |
| | ≥40 years old | 20 | 4 |
| **Sex identity** | Female | 499 | 99.6 |
| | Not-female | 1 | 0.2 |
| | No response | 1 | 0.2 |
| **Contraception type** | None | 447 | 89.2 |
| | Progestin-only | 31 | 6.2 |
| | Oestrogen & progestin | 23 | 4.6 |
| **Nulligravida** | Yes | 380 | 75.8 |
| | No | 121 | 24.2 |
| **Ethnicity** | White | 427 | 85.2 |
| | Mixed | 26 | 5.2 |
| | Asian / Asian British | 23 | 4.6 |
| | Black / Black British | 12 | 2.4 |
| | Other | 11 | 2.2 |
| | Middle Eastern | 2 | 0.4 |
| **Body mass index** | Healthy (18.5-24.9) | 238 | 47.5 |
| | Overweight (25.0-29.9) | 139 | 27.7 |
| | Obese (>30.0) | 113 | 22.6 |
| | Underweight (<18.5) | 11 | 2.2 |

**All variables are displayed as count (n) and percentage (%) of the total users (N = 501)**

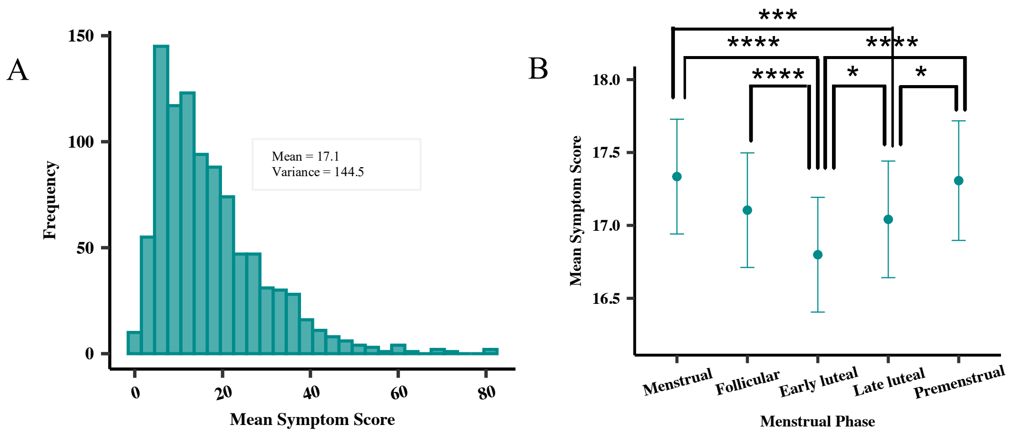

**Figure S8: Distribution of mean symptom score and how it varies over the menstrual cycle (disease cohort)**

(A) Distribution of mean symptom scores is shown as a histogram, binwidth = 0·5. (B) Mean symptom score was calculated by first summing the scores of each tracked symptom (all on a scale of 0-4) and secondly taking the mean across all days for each individual. Scatter plot shows mean symptom score split by menstrual phase, displayed as mean +/- SEM. The Friedman test plus Nemenyi pairwise comparisons was used to test the significance between mean symptom score and menstrual phase, due to the repeated measures nature of the data.

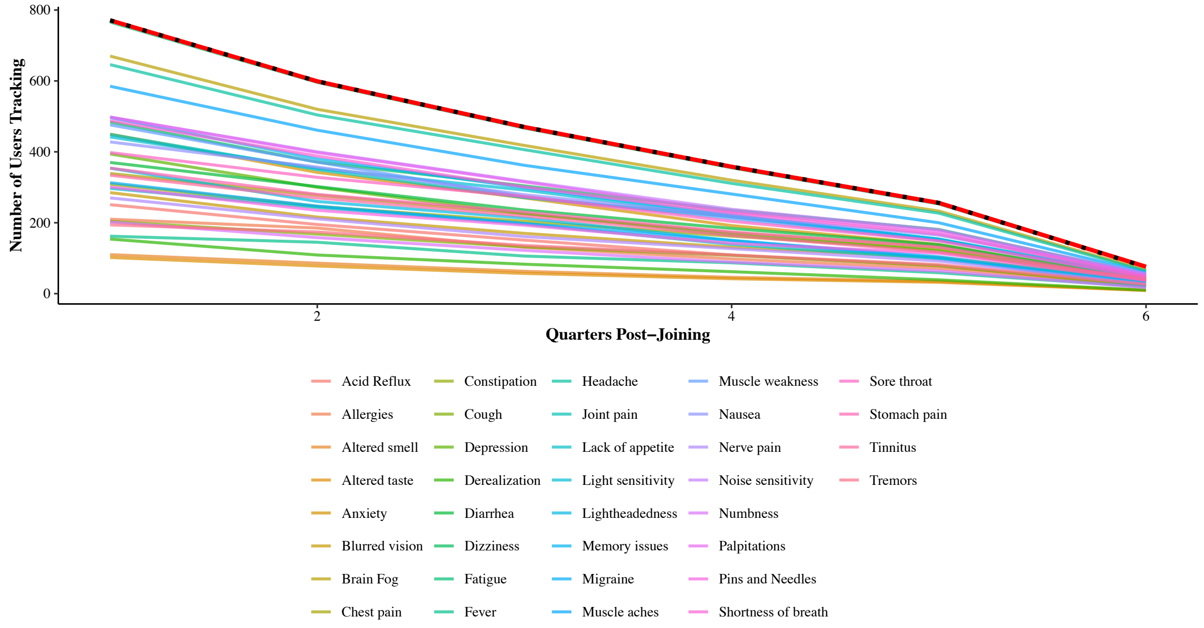

**Figure S9: Distribution of users tracking symptoms over study time period (disease cohort)**

The colored lines represent the number of users tracking each individual symptom per quarter since joining the study (symptoms=36). The x-axis shows quarters post-joining (Q1 = first quarter after a user joins), and the y-axis shows the number of users tracking that symptom. The bold red line indicates the total number of users logging their periods in each quarter. The dotted black line indicates the total number of users logging any symptom in each quarter.

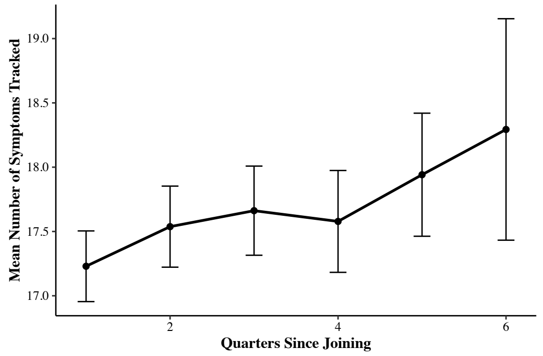

**Figure S10: The mean number of symptoms tracked over study time period (disease cohort)**
The x-axis shows quarters post-joining (Q1 = first quarter after a user joins), and the y-axis shows the mean number of symptoms tracked across all users.

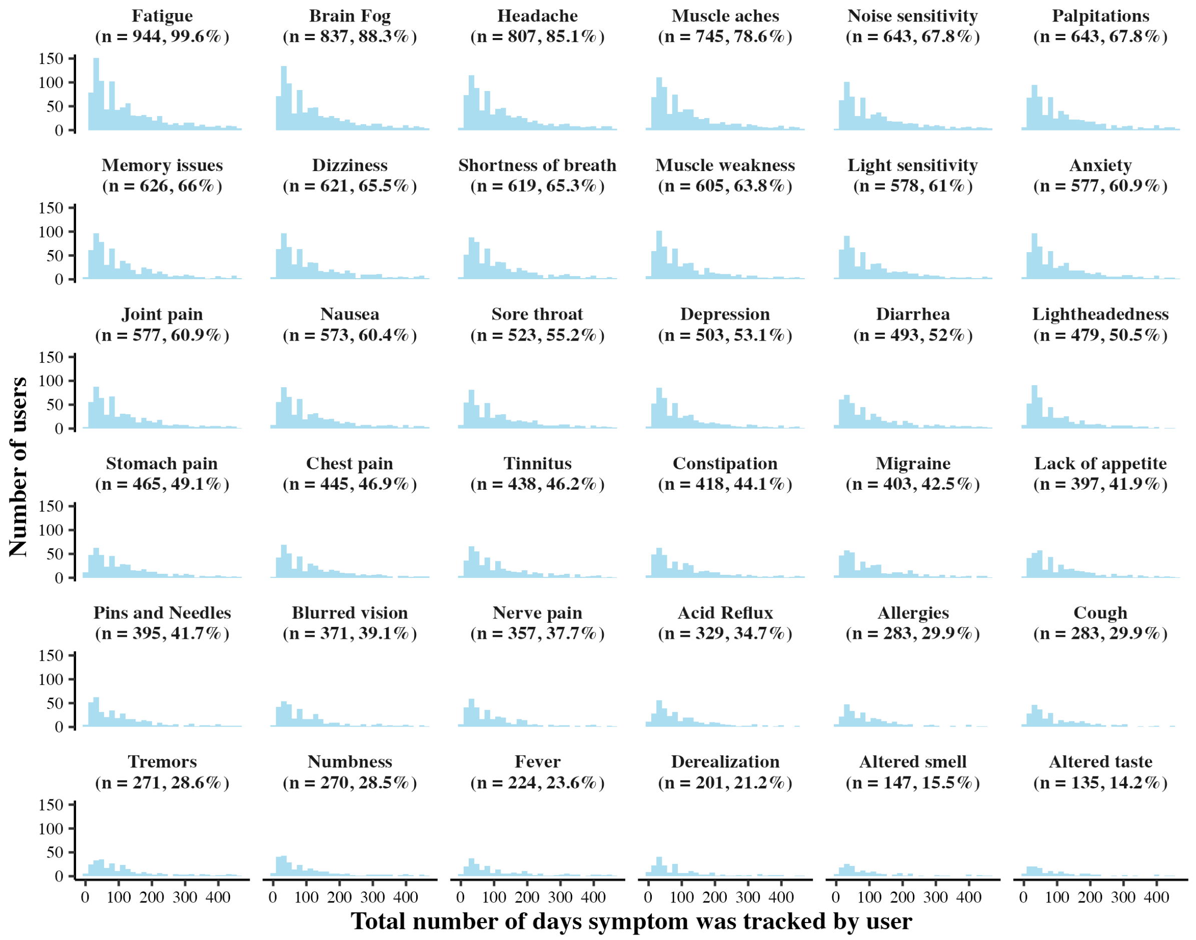

**Figure S11: Distribution of the number of days symptoms were tracked by users (disease cohort)**
The title of each facet contains the number and percentage of total users tracking that symptom at least once, facets are ordered from highest to lowest. Each histogram show the distribution of the total number of days each symptom was tracked by users.

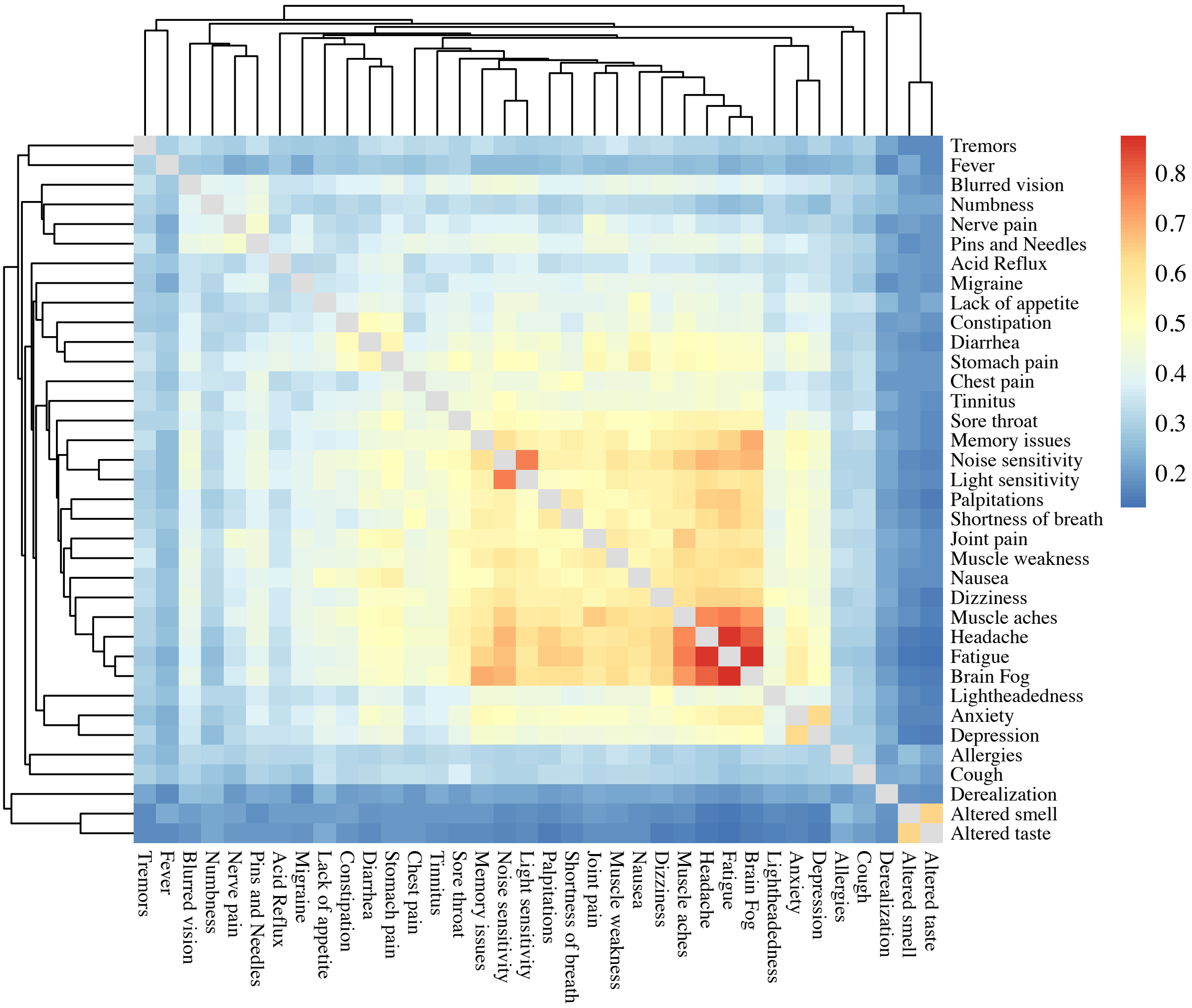

**Figure S12: Jaccard similarity heatmap of reported symptoms with hierarchical clustering (disease cohort)**

Heatmap visualizes the pairwise Jaccard similarity scores between 36 reported symptoms. The symptoms are listed along the x and y axes, with each cell representing the Jaccard similarity score between the symptoms corresponding to that row and column. The Jaccard similarity is a measure of similarity between two sets, defined as the size of the intersection divided by the size of the union of the sets of users reporting each pair of symptoms. Colours show the strength of being reported together, with scores close to 0 indicating the two symptoms rarely co-occur (blue) and scores close to 1 indicating the two symptoms frequently co-occur (red), n = 948. The grey cells represent comparisons of symptoms with themselves. Hierarchical clustering was performed on the Jaccard distances (1- Jaccard similarity) using the complete linkage method and resulting dendrograms are displayed along the secondary axes of the heatmap.

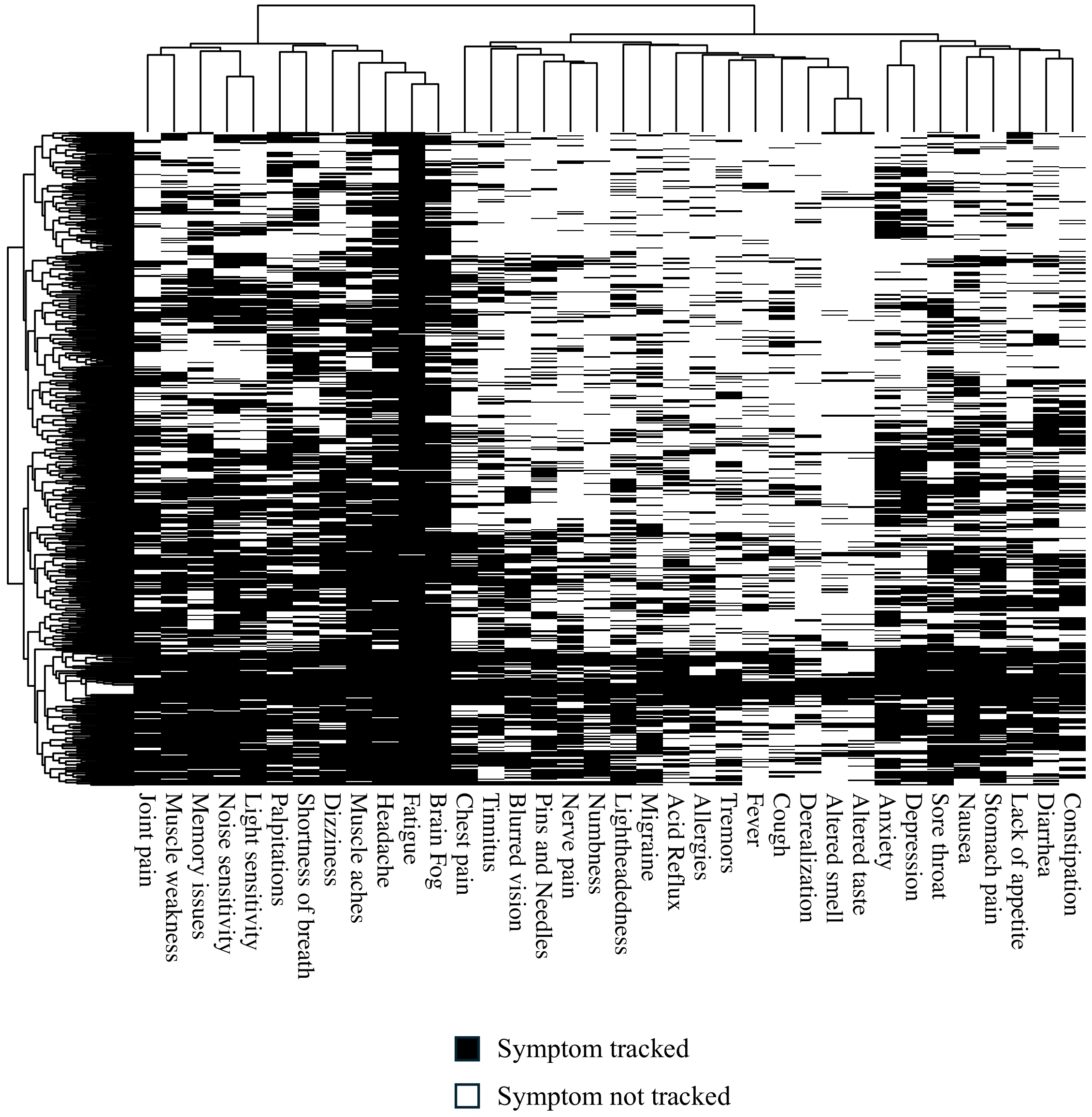

**Figure S13: Heatmap of symptoms and users with hierarchical clustering (disease cohort)**
Heatmap to visualize the presence (black) or absence (white) of symptoms tracked by users (N=948). Each cell in the heatmap corresponds to a user (y axis) – symptom (x axis) pair. Hierarchical clustering was performed using the complete linkage method and resulting dendrograms are displayed along the secondary axes of the heatmap.

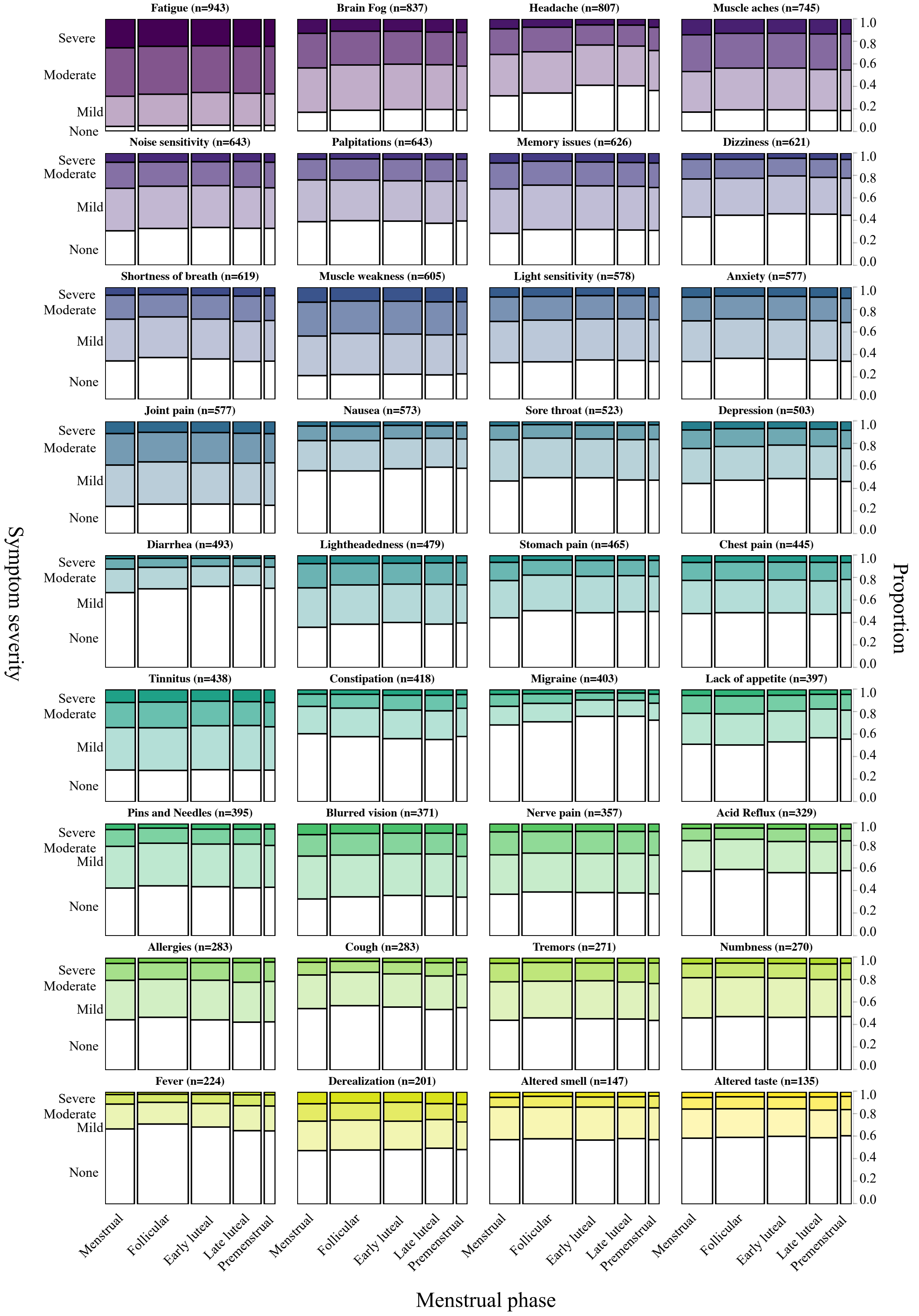

**Figure S14: Impact of menstrual cycle phase on individual symptom severity in the disease cohort**
Spineplots depict the frequencies in a two-way contingency table, with bar widths correspond to the relative frequencies of reports within a menstrual cycle phase and bar heights representing the conditional relative frequencies of the symptom severity rating in every menstrual cycle phase group.

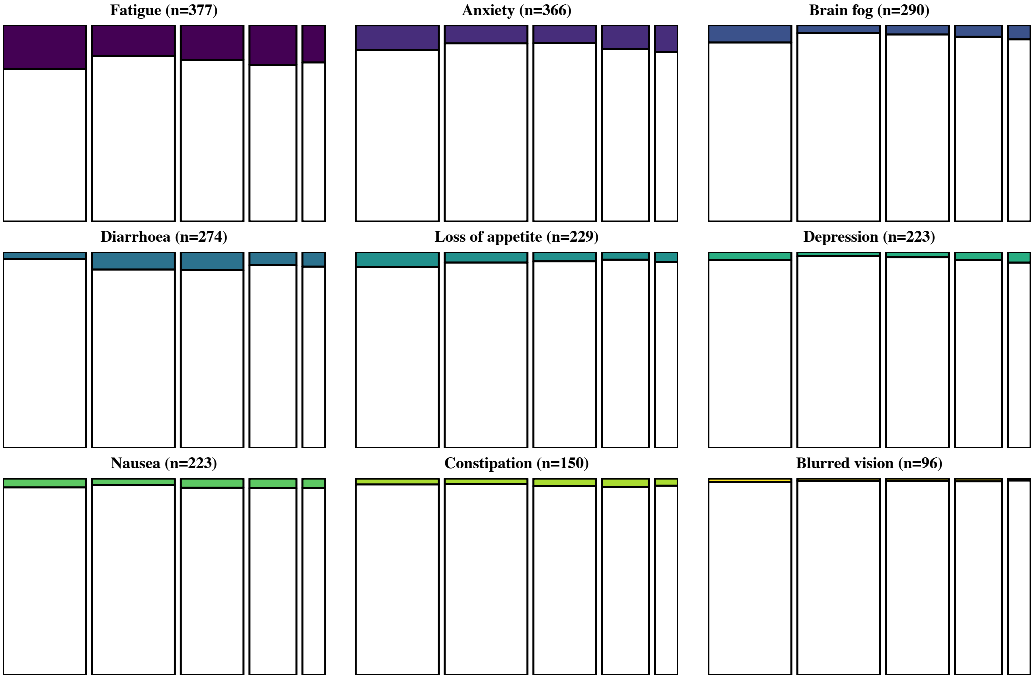

**Figure S15: Impact of menstrual cycle phase on individual symptom severity in the population cohort**
Spineplots depict the frequencies in a two-way contingency table, with bar widths correspond to the relative frequencies of reports within a menstrual cycle phase and bar heights representing the conditional relative frequencies of the symptom severity rating in every menstrual cycle phase group.

**Table S5: The fitting results of cumulative link mixed-effects models for individual symptoms joined disease and population cohorts**

**Table A. The fitting result of cumulative link mixed-effects model for fatigue (both cohorts)**

| **Fixed effects** | **Odds ratio (OR)** | **95% CI (lower)** | **95% CI (upper)** | **P value** |
|---|---|---|---|---|
| Menstrual phase | | | | |
| Menses | - | - | - | - |
| Follicular | 0.193 | 0.174 | 0.214 | **P < 0.001*** |
| Early luteal | 0.231 | 0.207 | 0.258 | **P < 0.001*** |
| Late luteal | 0.308 | 0.274 | 0.345 | **P < 0.001*** |
| Premenstrual | 0.391 | 0.336 | 0.455 | **P < 0.001*** |
| Cohort type | | | | |
| Population | - | - | - | - |
| Disease | 192 | 150 | 246 | **P < 0.001*** |
| Hormonal contraception type | | | | |
| None | - | - | - | - |
| Oestrogen & progestin | 1.04 | 0.717 | 1.52 | 0.824 |
| Progestin only | 1.19 | 0.877 | 1.63 | 0.261 |
| Age category | | | | |
| < 30 | - | - | - | - |
| 30-39 | 1.05 | 0.818 | 1.36 | 0.683 |
| >= 40 | 1.02 | 0.74 | 1.4 | 0.905 |
| Gravidity | | | | |
| Not-nulligravida | - | - | - | - |
| Nulligravida | 1.15 | 0.904 | 1.46 | 0.256 |
| Interaction of cohort type and menstrual phase | | | | |
| Disease_cohort:Menstrual_phase_Menses | - | - | - | - |
| Disease_cohort:Menstrual_phase_Follicular | 4.95 | 4.44 | 5.53 | **P < 0.001*** |
| Disease_cohort:Menstrual_phaseEarly luteal | 3.82 | 3.41 | 4.28 | **P < 0.001*** |
| Disease_cohort:Menstrual_phaseLate luteal | 3 | 2.66 | 3.39 | **P < 0.001*** |
| Disease_cohort:Menstrual_phasePremenstrual | 2.38 | 2.03 | 2.78 | **P < 0.001*** |

* P value <0·05

**Table B. The fitting result of cumulative link mixed-effects model for brain fog (both cohorts)**

| **Fixed effects** | **Odds ratio (OR)** | **95% CI (lower)** | **95% CI (upper)** | **P value** |
|---|---|---|---|---|
| Menstrual phase | | | | |
| Menses | - | - | - | - |
| Follicular | 0.192 | 0.162 | 0.226 | **P < 0.001*** |
| Early luteal | 0.225 | 0.19 | 0.267 | **P < 0.001*** |
| Late luteal | 0.307 | 0.258 | 0.367 | **P < 0.001*** |
| Premenstrual | 0.496 | 0.398 | 0.617 | **P < 0.001*** |
| Cohort type | | | | |
| Population | - | - | - | - |
| Disease | 196 | 142 | 270 | **P < 0.001*** |
| Hormonal contraception type | | | | |
| None | - | - | - | - |
| Oestrogen & progestin | 0.569 | 0.356 | 0.911 | **0.019*** |
| Progestin only | 0.88 | 0.577 | 1.34 | 0.552 |
| Age category | | | | |
| < 30 | - | - | - | - |
| 30-39 | 0.866 | 0.622 | 1.21 | 0.396 |
| >= 40 | 0.708 | 0.476 | 1.05 | 0.087 |
| Gravidity | | | | |
| Not-nulligravida | - | - | - | - |
| Nulligravida | 1.02 | 0.756 | 1.37 | 0.912 |
| Interaction of cohort type and menstrual phase | | | | |
| Disease_cohort:Menstrual_phase_Menses | - | - | - | - |
| Disease_cohort:Menstrual_phase_Follicular | 4.97 | 4.2 | 5.88 | **P < 0.001*** |
| Disease_cohort:Menstrual_phaseEarly luteal | 3.86 | 3.24 | 4.6 | **P < 0.001*** |
| Disease_cohort:Menstrual_phaseLate luteal | 2.91 | 2.43 | 3.49 | **P < 0.001*** |
| Disease_cohort:Menstrual_phasePremenstrual | 1.92 | 1.53 | 2.4 | **P < 0.001*** |

* P value <0·05

**Table C. The fitting result of cumulative link mixed-effects model for headache (both cohorts)**

| Fixed effects | Odds ratio (OR) | 95% CI (lower) | 95% CI (upper) | P value |
|---|---|---|---|---|
| Menstrual phase | | | | |
| Menses | - | - | - | - |
| Follicular | 0.314 | 0.281 | 0.35 | **P < 0.001*** |
| Early luteal | 0.217 | 0.191 | 0.246 | **P < 0.001*** |
| Late luteal | 0.255 | 0.222 | 0.292 | **P < 0.001*** |
| Premenstrual | 0.418 | 0.352 | 0.495 | **P < 0.001*** |
| Cohort type | | | | |
| Population | - | - | - | - |
| Disease | 22.7 | 17.9 | 28.9 | **P < 0.001*** |
| Hormonal contraception type | | | | |
| None | - | - | - | - |
| Oestrogen & progestin | 1.06 | 0.723 | 1.55 | 0.770 |
| Progestin only | 1.08 | 0.782 | 1.5 | 0.634 |
| Age category | | | | |
| < 30 | - | - | - | - |
| 30-39 | 1.09 | 0.846 | 1.4 | 0.516 |
| >= 40 | 0.865 | 0.631 | 1.19 | 0.371 |
| Gravidity | | | | |
| Not-nulligravida | - | - | - | - |
| Nulligravida | 0.975 | 0.766 | 1.24 | 0.838 |
| Interaction of cohort type and menstrual phase | | | | |
| Disease_cohort:Menstrual_phase_Menses | - | - | - | - |
| Disease_cohort:Menstrual_phase_Follicular | 2.82 | 2.51 | 3.16 | **P < 0.001*** |
| Disease_cohort:Menstrual_phaseEarly luteal | 2.77 | 2.43 | 3.17 | **P < 0.001*** |
| Disease_cohort:Menstrual_phaseLate luteal | 2.46 | 2.13 | 2.84 | **P < 0.001*** |
| Disease_cohort:Menstrual_phasePremenstrual | 1.95 | 1.63 | 2.33 | **P < 0.001*** |

* P value <0·05

**Table S6: The fitting results of cumulative link mixed-effects models for individual symptoms in the disease cohort**

| **Table A. The fitting results of cumulative link mixed-effects model for fatigue (disease cohort)** | | | |
|---|---|---|---|
| AIC: 212000 BIC: 212000 Log-Likelihood: -106000 Number of Observations: 119203 Number of Users: 943 | | | |
| **Random effect** | **Variance** | **Standard deviation** | |
| Individual (intercept) | 3.995 | 1.999 | |
| **Fixed effects** | **Estimate** | **Standard error** | **P value** |
| Follicular | -0.0548 | 0.0174 | 1.7E-03* |
| Early luteal | -0.151 | 0.0183 | 1.47E-16* |
| Late luteal | -0.0969 | 0.0197 | 9.07E-07* |
| Premenstrual | -0.0883 | 0.0264 | 8.22E-04* |
| **Threshold coefficients** | **Estimate** | **Standard error** | **Z value** |
| 1 (Threshold 0 to 1) | -4.64 | 0.0692 | -67 |
| 2 (Threshold 1 to 2) | -1.26 | 0.0668 | -18.9 |
| 3 (Threshold 2 to 3) | 1.64 | 0.0669 | 24.5 |

| **Table B. The fitting results of cumulative link mixed-effects model for brain fog (disease cohort)** | | | |
|---|---|---|---|
| AIC: 184000 BIC: 184000 Log-Likelihood: -91900 Number of Observations: 104753 Number of Users: 837 | | | |
| **Random effect** | **Variance** | **Standard deviation** | |
| Individual (intercept) | 6.173 | 2.485 | |
| **Fixed effects** | **Estimate** | **Standard error** | **P value** |
| Follicular | -0.059 | 0.0188 | 1.73E-03* |
| Early luteal | -0.161 | 0.0197 | 2.57E-16* |
| Late luteal | -0.131 | 0.0213 | 7.66E-10* |
| Premenstrual | -0.0635 | 0.0284 | 2.55E-02* |
| **Threshold coefficients** | **Estimate** | **Standard error** | **Z value** |
| 1 (Threshold 0 to 1) | -2.71 | 0.0898 | -30.2 |
| 2 (Threshold 1 to 2) | 0.482 | 0.0894 | 5.39 |
| 3 (Threshold 2 to 3) | 3.46 | 0.0903 | 38.3 |

| **Table C. The fitting results of cumulative link mixed-effects model for headache (disease cohort)** | | | |
|---|---|---|---|
| AIC: 197000 BIC: 197000 Log-Likelihood: -98700 Number of Observations: 102456 Number of Users: 807 | | | |
| **Random effect** | **Variance** | **Standard deviation** | |
| Individual (intercept) | 3.047 | 1.746 | |
| **Fixed effects** | **Estimate** | **Standard error** | **P value** |
| Follicular | -0.149 | 0.0183 | 4.12E-16* |
| Early luteal | -0.591 | 0.0194 | 1.19E-204* |
| Late luteal | -0.543 | 0.0209 | 5.4E-148* |
| Premenstrual | -0.249 | 0.0278 | 2.85E-19* |
| **Threshold coefficients** | **Estimate** | **Standard error** | **Z value** |
| 1 (Threshold 0 to 1) | -1.28 | 0.0638 | -20 |
| 2 (Threshold 1 to 2) | 1.11 | 0.0638 | 17.4 |
| 3 (Threshold 2 to 3) | 3.46 | 0.0652 | 53 |

| Table D. The fitting results of cumulative link mixed-effects model for muscle aches (disease cohort) | | | |
|---|---|---|---|
| AIC: 164000 BIC: 164000 Log-Likelihood: -81800 Number of Observations: 91200 Number of Users: 745 | | | |
| **Random effect** | **Variance** | **Standard deviation** | |
| Individual (intercept) | 6.342 | 2.518 | |
| **Fixed effects** | **Estimate** | **Standard error** | **P value** |
| Follicular | -0.112 | 0.02 | 2.48E-08* |
| Early luteal | -0.162 | 0.0209 | 8.41E-15* |
| Late luteal | -0.0748 | 0.0226 | 9.2E-04* |
| Premenstrual | -0.0573 | 0.03 | 5.65E-02 |
| **Threshold coefficients** | **Estimate** | **Standard error** | **Z value** |
| 1 (Threshold 0 to 1) | -2.76 | 0.0942 | -29.3 |
| 2 (Threshold 1 to 2) | 0.288 | 0.0937 | 3.07 |
| 3 (Threshold 2 to 3) | 3.19 | 0.0944 | 33.8 |

| Table E. The fitting results of cumulative link mixed-effects model for noise sensitivity (disease cohort) | | | |
|---|---|---|---|
| AIC: 131000 BIC: 131000 Log-Likelihood: -65300 Number of Observations: 79515 Number of Users: 643 | | | |
| **Random effect** | **Variance** | **Standard deviation** | |
| Individual (intercept) | 7.813 | 2.795 | |
| **Fixed effects** | **Estimate** | **Standard error** | **P value** |
| Follicular | -0.0416 | 0.0222 | 6.02E-02 |
| Early luteal | -0.133 | 0.0232 | 9.86E-09* |
| Late luteal | -0.0768 | 0.0251 | 2.21E-03* |
| Premenstrual | -0.0481 | 0.0337 | 1.53E-01 |
| **Threshold coefficients** | **Estimate** | **Standard error** | **Z value** |
| 1 (Threshold 0 to 1) | -1.51 | 0.0712 | -21.3 |
| 2 (Threshold 1 to 2) | 1.58 | 0.0707 | 22.4 |
| 3 (Threshold 2 to 3) | 4.47 | 0.0707 | 63.3 |

| Table F. The fitting results of cumulative link mixed-effects model for palpitations (disease cohort) | | | |
|---|---|---|---|
| AIC: 133000 BIC: 133000 Log-Likelihood: -66300 Number of Observations: 77589 Number of Users: 643 | | | |
| **Random effect** | **Variance** | **Standard deviation** | |
| Individual (intercept) | 5.205 | 2.281 | |
| **Fixed effects** | **Estimate** | **Standard error** | **P value** |
| Follicular | 0.0486 | 0.0173 | 4.94E-03* |
| Early luteal | 0.081 | 0.018 | 6.43E-06* |
| Late luteal | 0.172 | 0.0192 | 2.54E-19* |
| Premenstrual | 0.0823 | 0.0239 | 5.67E-04* |
| **Threshold coefficients** | **Estimate** | **Standard error** | **Z value** |
| 1 (Threshold 0 to 1) | -0.76 | 0.0233 | -32.7 |
| 2 (Threshold 1 to 2) | 2.01 | 0.0222 | 90.5 |
| 3 (Threshold 2 to 3) | 4.56 | 0.0248 | 184 |

| Table G. The fitting results of cumulative link mixed-effects model for memory issues (disease cohort) | | | |
|---|---|---|---|
| AIC: 112000 BIC: 112000 Log-Likelihood: -56100 Number of Observations: 74662 Number of | | | |
| **Random effect** | **Variance** | **Standard deviation** | |
| Individual (intercept) | 9.18 | 3.03 | |
| **Fixed effects** | **Estimate** | **Standard error** | **P value** |
| Follicular | -0.0847 | 0.00386 | 1.56E-106* |
| Early luteal | -0.123 | 0.00411 | 3.99E-197* |
| Late luteal | -0.0675 | 0.00404 | 1.54E-62* |
| Premenstrual | 0.00249 | 0.00407 | 5.41E-01 |
| **Threshold coefficients** | **Estimate** | **Standard error** | **Z value** |
| 1 (Threshold 0 to 1) | -1.92 | 0.00402 | -478 |
| 2 (Threshold 1 to 2) | 1.57 | 0.00254 | 620 |
| 3 (Threshold 2 to 3) | 4.69 | 0.00451 | 1040 |

| Table H. The fitting results of cumulative link mixed-effects model for dizziness (disease cohort) | | | |
|---|---|---|---|
| AIC: 126000 BIC: 126000 Log-Likelihood: -62900 Number of Observations: 76626 Number of Users: 622 | | | |
| **Random effect** | **Variance** | **Standard deviation** | |
| Individual (intercept) | 5.159 | 2.271 | |
| **Fixed effects** | **Estimate** | **Standard error** | **P value** |
| Follicular | 0.00344 | 0.023 | 8.81E-01 |
| Early luteal | -0.171 | 0.0241 | 1.12E-12* |
| Late luteal | -0.092 | 0.026 | 4.02E-04* |
| Premenstrual | -0.0348 | 0.0346 | 3.14E-01 |
| **Threshold coefficients** | **Estimate** | **Standard error** | **Z value** |
| 1 (Threshold 0 to 1) | -0.524 | 0.0946 | -5.54 |
| 2 (Threshold 1 to 2) | 2.11 | 0.0949 | 22.3 |
| 3 (Threshold 2 to 3) | 4.46 | 0.0967 | 46.2 |

| Table I. The fitting results of cumulative link mixed-effects model for shortness of breath (disease | | | |
|---|---|---|---|
| AIC: 129000 BIC: 129000 Log-Likelihood: -64700 Number of Observations: 75074 Number of Users: 619 | | | |
| **Random effect** | **Variance** | **Standard deviation** | |
| Individual (intercept) | 5.729 | 2.394 | |
| **Fixed effects** | **Estimate** | **Standard error** | **P value** |
| Follicular | -0.079 | 0.0225 | 4.58E-04* |
| Early luteal | 0.00656 | 0.0235 | 7.8E-01 |
| Late luteal | 0.171 | 0.0254 | 1.61E-11* |
| Premenstrual | 0.127 | 0.0339 | 1.69E-04* |
| **Threshold coefficients** | **Estimate** | **Standard error** | **Z value** |
| 1 (Threshold 0 to 1) | -0.922 | 0.0993 | -9.28 |
| 2 (Threshold 1 to 2) | 1.86 | 0.0995 | 18.7 |
| 3 (Threshold 2 to 3) | 4.53 | 0.101 | 44.8 |

| Table J. The fitting results of cumulative link mixed-effects model for muscle weakness (disease cohort) | | | |
|---|---|---|---|
| AIC: 124000 BIC: 124000 Log-Likelihood: -62000 Number of Observations: 70122 Number of Users: 605 | | | |
| **Random effect** | **Variance** | **Standard deviation** | |
| Individual (intercept) | 6.915 | 2.63 | |
| **Fixed effects** | **Estimate** | **Standard error** | **P value** |
| Follicular | -0.0656 | 0.023 | 4.31E-03* |
| Early luteal | -0.0846 | 0.024 | 4.22E-04* |
| Late luteal | -0.0194 | 0.0259 | 4.55E-01 |
| Premenstrual | -0.0728 | 0.0346 | 3.55E-02* |
| **Threshold coefficients** | **Estimate** | **Standard error** | **Z value** |
| 1 (Threshold 0 to 1) | -2.24 | 0.11 | -20.4 |
| 2 (Threshold 1 to 2) | 0.758 | 0.11 | 6.9 |
| 3 (Threshold 2 to 3) | 3.69 | 0.111 | 33.3 |

| Table K. The fitting results of cumulative link mixed-effects model for light sensitivity (disease cohort) | | | |
|---|---|---|---|
| AIC: 112000 BIC: 112000 Log-Likelihood: -56100 Number of Observations: 69286 Number of | | | |
| Random effect | Variance | Standard deviation | |
| Individual (intercept) | 7.688 | 2.773 | |
| **Fixed effects** | **Estimate** | **Standard error** | **P value** |
| Follicular | -0.0245 | 0.024 | 3.08E-01 |
| Early luteal | -0.165 | 0.0252 | 5.53E-11* |
| Late luteal | -0.159 | 0.0272 | 4.85E-09* |
| Premenstrual | -0.0829 | 0.0364 | 2.27E-02* |
| **Threshold coefficients** | **Estimate** | **Standard error** | **Z value** |
| 1 (Threshold 0 to 1) | -1.4 | 0.118 | -11.8 |
| 2 (Threshold 1 to 2) | 1.71 | 0.119 | 14.4 |
| 3 (Threshold 2 to 3) | 4.52 | 0.12 | 37.6 |

| Table L. The fitting results of cumulative link mixed-effects model for anxiety (disease cohort) | | | |
|---|---|---|---|
| AIC: 123000 BIC: 123000 Log-Likelihood: -61500 Number of Observations: 66424 Number of Users: | | | |
| **Random effect** | **Variance** | **Standard deviation** | |
| Individual (intercept) | 4.445 | 2.108 | |
| **Fixed effects** | **Estimate** | **Standard error** | **P value** |
| Follicular | -0.0222 | 0.0234 | 3.42E-01 |
| Early luteal | -0.0442 | 0.0245 | 7.11E-02 |
| Late luteal | 0.0317 | 0.0264 | 2.31E-01 |
| Premenstrual | 0.13 | 0.0354 | 2.38E-04* |
| **Threshold coefficients** | **Estimate** | **Standard error** | **Z value** |
| 1 (Threshold 0 to 1) | -1.07 | 0.0906 | -11.8 |
| 2 (Threshold 1 to 2) | 1.44 | 0.0907 | 15.9 |
| 3 (Threshold 2 to 3) | 3.71 | 0.0921 | 40.2 |

| Table M. The fitting results of cumulative link mixed-effects model for joint pain (disease cohort) | | | |
|---|---|---|---|
| AIC: 118000 BIC: 118000 Log-Likelihood: -59200 Number of Observations: 68637 Number of | | | |
| **Random effect** | **Variance** | **Standard deviation** | |
| Individual (intercept) | 7.45 | 2.73 | |
| **Fixed effects** | **Estimate** | **Standard error** | **P value** |
| Follicular | -0.0779 | 0.0239 | 1.11E-03* |
| Early luteal | -0.0969 | 0.0247 | 8.93E-05* |
| Late luteal | -0.0501 | 0.0267 | 6.1E-02 |
| Premenstrual | -0.0108 | 0.0355 | 7.61E-01 |
| **Threshold coefficients** | **Estimate** | **Standard error** | **Z value** |
| 1 (Threshold 0 to 1) | -2.21 | 0.128 | -17.2 |
| 2 (Threshold 1 to 2) | 0.814 | 0.128 | 6.35 |
| 3 (Threshold 2 to 3) | 3.72 | 0.13 | 28.7 |

| Table N. The fitting results of cumulative link mixed-effects model for nausea (disease cohort) | | | |
|---|---|---|---|
| AIC: 112000 BIC: 112000 Log-Likelihood: -56000 Number of Observations: 69149 Number of Users: 576 | | | |
| **Random effect** | **Variance** | **Standard deviation** | |
| Individual (intercept) | 3.39 | 1.841 | |
| **Fixed effects** | **Estimate** | **Standard error** | **P value** |
| Follicular | 0.0375 | 0.0245 | 1.25E-01 |
| Early luteal | -0.0976 | 0.0257 | 1.47E-04* |
| Late luteal | -0.151 | 0.0279 | 6.34E-08* |
| Premenstrual | -0.0831 | 0.0372 | 2.56E-02* |
| **Threshold coefficients** | **Estimate** | **Standard error** | **Z value** |
| 1 (Threshold 0 to 1) | 0.303 | 0.08 | 3.79 |
| 2 (Threshold 1 to 2) | 2.41 | 0.0806 | 29.9 |
| 3 (Threshold 2 to 3) | 4.57 | 0.0835 | 54.7 |

| Table O. The fitting results of cumulative link mixed-effects model for sore throat (disease cohort) | | | |
|---|---|---|---|
| AIC: 102000 BIC: 102000 Log-Likelihood: -50900 Number of Observations: 64314 Number of Users: 524 | | | |
| **Random effect** | **Variance** | **Standard deviation** | |
| Individual (intercept) | 3.845 | 1.961 | |
| **Fixed effects** | **Estimate** | **Standard error** | **P value** |
| Follicular | -0.064 | 0.0253 | 1.14E-02* |
| Early luteal | -0.0915 | 0.0264 | 5.37E-04* |
| Late luteal | 0.026 | 0.0284 | 3.59E-01 |
| Premenstrual | 0.0248 | 0.0378 | 5.12E-01 |
| **Threshold coefficients** | **Estimate** | **Standard error** | **Z value** |
| 1 (Threshold 0 to 1) | -0.339 | 0.0894 | -3.8 |
| 2 (Threshold 1 to 2) | 2.44 | 0.0901 | 27.1 |
| 3 (Threshold 2 to 3) | 5.02 | 0.0946 | 53.1 |

| Table P. The fitting results of cumulative link mixed-effects model for depression (disease cohort) | | | |
|---|---|---|---|
| AIC: 95500 BIC: 95600 Log-Likelihood: -47800 Number of Observations: 55988 Number of Users: 503 * | | | |
| **Random effect** | **Variance** | **Standard deviation** | |
| Individual (intercept) | 4.576 | 2.139 | |
| **Fixed effects** | **Estimate** | **Standard error** | **P value** |
| Follicular | -0.0959 | 0.0267 | 3.21E-04* |
| Early luteal | -0.231 | 0.028 | 1.58E-16* |
| Late luteal | -0.164 | 0.0303 | 6.12E-08* |
| Premenstrual | 0.00814 | 0.0402 | 8.4E-01 |
| **Threshold coefficients** | **Estimate** | **Standard error** | **Z value** |
| 1 (Threshold 0 to 1) | -0.424 | 0.0984 | -4.31 |
| 2 (Threshold 1 to 2) | 1.87 | 0.0988 | 18.9 |
| 3 (Threshold 2 to 3) | 4.01 | 0.101 | 39.8 |

| Table Q. The fitting results of cumulative link mixed-effects model for diarrhoea (disease cohort) | | | |
|---|---|---|---|
| AIC: 82500 BIC: 82500 Log-Likelihood: -41200 Number of Observations: 58432 Number of Users: 493 * | | | |
| **Random effect** | **Variance** | **Standard deviation** | |
| Individual (intercept) | 2.485 | 1.577 | |
| **Fixed effects** | **Estimate** | **Standard error** | **P value** |
| Follicular | -0.183 | 0.0286 | 1.52E-10* |
| Early luteal | -0.318 | 0.0302 | 7.07E-26* |
| Late luteal | -0.366 | 0.033 | 1.76E-28* |
| Premenstrual | -0.209 | 0.0438 | 1.8E-06* |
| **Threshold coefficients** | **Estimate** | **Standard error** | **Z value** |
| 1 (Threshold 0 to 1) | 0.997 | 0.0764 | 13.1 |
| 2 (Threshold 1 to 2) | 2.68 | 0.0773 | 34.6 |
| 3 (Threshold 2 to 3) | 4.48 | 0.0815 | 54.9 |

| Table R. The fitting results of cumulative link mixed-effects model for lightheadedness (disease cohort) | | | |
|---|---|---|---|
| AIC: 82000 BIC:82100 Log-Likelihood: -41000 Number of Observations: 48662 Number of Users: 479 * | | | |
| **Random effect** | **Variance** | **Standard deviation** | |
| Individual (intercept) | 6.098 | 2.469 | |
| **Fixed effects** | **Estimate** | **Standard error** | **P value** |
| Follicular | -0.0784 | 0.0284 | 5.87E-03* |
| Early luteal | -0.196 | 0.0296 | 3.42E-11* |
| Late luteal | -0.136 | 0.0319 | 1.93E-05* |
| Premenstrual | -0.164 | 0.0427 | 1.18E-04* |
| **Threshold coefficients** | **Estimate** | **Standard error** | **Z value** |
| 1 (Threshold 0 to 1) | -0.954 | 0.117 | -8.15 |
| 2 (Threshold 1 to 2) | 1.81 | 0.117 | 15.4 |
| 3 (Threshold 2 to 3) | 4.37 | 0.119 | 36.6 |

| Table S. The fitting results of cumulative link mixed-effects model for stomach pain (disease cohort) | | | |
|---|---|---|---|
| AIC: 93600 BIC: 93700 Log-Likelihood: -46800 Number of Observations: 55377 Number of Users: 466 * | | | |
| **Random effect** | **Variance** | **Standard deviation** | |
| Individual (intercept) | 3.921 | 1.98 | |
| **Fixed effects** | **Estimate** | **Standard error** | **P value** |
| Follicular | -0.382 | 0.0266 | 9.57E-47* |
| Early luteal | -0.294 | 0.0277 | 3.04E-26* |
| Late luteal | -0.35 | 0.0301 | 2.84E-31* |
| Premenstrual | -0.336 | 0.0402 | 6.92E-17* |
| **Threshold coefficients** | **Estimate** | **Standard error** | **Z value** |
| 1 (Threshold 0 to 1) | -0.415 | 0.095 | -4.37 |
| 2 (Threshold 1 to 2) | 1.99 | 0.0955 | 20.9 |
| 3 (Threshold 2 to 3) | 4.24 | 0.0981 | 43.2 |

| Table T. The fitting results of cumulative link mixed-effects model for chest pain (disease cohort) | | | |
|---|---|---|---|
| AIC: 83000 BIC: 83000 Log-Likelihood: -41500 Number of Observations: 54259 Number of Users: 447 * | | | |
| **Random effect** | **Variance** | **Standard deviation** | |
| Individual (intercept) | 6.184 | 2.487 | |
| **Fixed effects** | **Estimate** | **Standard error** | **P value** |
| Follicular | 0.0519 | 0.0283 | 6.65E-02 |
| Early luteal | -0.000598 | 0.0296 | 9.84E-01 |
| Late luteal | 0.108 | 0.0319 | 6.87E-04* |
| Premenstrual | 0.028 | 0.0428 | 5.12E-01 |
| **Threshold coefficients** | **Estimate** | **Standard error** | **Z value** |
| 1 (Threshold 0 to 1) | -0.211 | 0.121 | -1.75 |
| 2 (Threshold 1 to 2) | 2.39 | 0.121 | 19.7 |
| 3 (Threshold 2 to 3) | 4.94 | 0.124 | 39.9 |

| Table U. The fitting results of cumulative link mixed-effects model for tinnitus (disease cohort) | | | |
|---|---|---|---|
| AIC: 8500 BIC: 78600 Log-Likelihood: -39300 Number of Observations: 52587 Number of Users: 438 * | | | |
| **Random effect** | **Variance** | **Standard deviation** | |
| Individual (intercept) | 11.49 | 3.39 | |
| **Fixed effects** | **Estimate** | **Standard error** | **P value** |
| Follicular | -0.0138 | 0.0285 | 6.28E-01 |
| Early luteal | -0.0874 | 0.0298 | 3.33E-03* |
| Late luteal | -0.0637 | 0.0321 | 4.72E-02* |
| Premenstrual | -0.0161 | 0.0429 | 7.08E-01 |
| **Threshold coefficients** | **Estimate** | **Standard error** | **Z value** |
| 1 (Threshold 0 to 1) | -1.97 | 0.164 | -12 |
| 2 (Threshold 1 to 2) | 1.64 | 0.164 | 9.99 |
| 3 (Threshold 2 to 3) | 4.81 | 0.166 | 29.1 |

| Table V. The fitting results of cumulative link mixed-effects model for constipation (disease cohort) | | | |
|---|---|---|---|
| AIC: 81800 BIC: 81900 Log-Likelihood: -40900 Number of Observations: 48728 Number of Users: 418 | | | |
| **Random effect** | **Variance** | **Standard deviation** | |
| Individual (intercept) | 3.545 | 1.883 | |
| **Fixed effects** | **Estimate** | **Standard error** | **P value** |
| Follicular | 0.137 | 0.0295 | 3.24E-06* |
| Early luteal | 0.297 | 0.0306 | 3.31E-22* |
| Late luteal | 0.354 | 0.0329 | 5.91E-27* |
| Premenstrual | 0.165 | 0.0441 | 1.91E-04* |
| **Threshold coefficients** | **Estimate** | **Standard error** | **Z value** |
| 1 (Threshold 0 to 1) | 0.587 | 0.0965 | 6.08 |
| 2 (Threshold 1 to 2) | 2.46 | 0.0972 | 25.3 |
| 3 (Threshold 2 to 3) | 4.45 | 0.1 | 44.5 |

| Table W. The fitting results of cumulative link mixed-effects model for migraine (disease cohort) | | | |
|---|---|---|---|
| AIC: 60500 BIC:60600 Log-Likelihood: -30200 Number of Observations: 46884 Number of Users: 403 * | | | |
| **Random effect** | **Variance** | **Standard deviation** | |
| Individual (intercept) | 3.668 | 1.915 | |
| **Fixed effects** | **Estimate** | **Standard error** | **P value** |
| Follicular | -0.182 | 0.0332 | 4.18E-08* |
| Early luteal | -0.585 | 0.0359 | 1.34E-59* |
| Late luteal | -0.582 | 0.0392 | 8.6E-50* |
| Premenstrual | -0.297 | 0.0514 | 7.9E-09* |
| **Threshold coefficients** | **Estimate** | **Standard error** | **Z value** |
| 1 (Threshold 0 to 1) | 1.3 | 0.103 | 12.7 |
| 2 (Threshold 1 to 2) | 2.84 | 0.104 | 27.5 |
| 3 (Threshold 2 to 3) | 4.58 | 0.107 | 42.8 |

| Table X. The fitting results of cumulative link mixed-effects model for lack of appetite (disease cohort) | | | |
|---|---|---|---|
| AIC: 73900 BIC: 74000 Log-Likelihood: -37000 Number of Observations: 46147 Number of Users: 397 | | | |
| **Random effect** | **Variance** | **Standard deviation** | |
| Individual (intercept) | 4.801 | 2.191 | |
| **Fixed effects** | **Estimate** | **Standard error** | **P value** |
| Follicular | 0.126 | 0.0299 | 2.47E-05* |
| Early luteal | -0.0996 | 0.0315 | 1.59E-03* |
| Late luteal | -0.329 | 0.0345 | 1.35E-21* |
| Premenstrual | -0.257 | 0.0461 | 2.41E-08* |
| **Threshold coefficients** | **Estimate** | **Standard error** | **Z value** |
| 1 (Threshold 0 to 1) | -0.0208 | 0.114 | -0.183 |
| 2 (Threshold 1 to 2) | 2.21 | 0.115 | 19.3 |
| 3 (Threshold 2 to 3) | 4.57 | 0.117 | 38.9 |

| Table Y. The fitting results of cumulative link mixed-effects model for pins and needles (disease cohort) | | | |
|---|---|---|---|
| AIC: 71600 BIC: 71700 Log-Likelihood: -35800 Number of Observations: 47249 Number of Users: 395 * | | | |
| **Random effect** | **Variance** | **Standard deviation** | |
| Individual (intercept) | 7.005 | 2.647 | |
| **Fixed effects** | **Estimate** | **Standard error** | **P value** |
| Follicular | -0.035 | 0.0299 | 2.43E-01 |
| Early luteal | -0.0517 | 0.0314 | 9.91E-02 |
| Late luteal | -0.0133 | 0.0338 | 6.94E-01 |
| Premenstrual | 0.019 | 0.0451 | 6.73E-01 |
| **Threshold coefficients** | **Estimate** | **Standard error** | **Z value** |
| 1 (Threshold 0 to 1) | -0.292 | 0.137 | -2.13 |
| 2 (Threshold 1 to 2) | 2.91 | 0.138 | 21.1 |
| 3 (Threshold 2 to 3) | 5.43 | 0.141 | 38.5 |

| Table Z. The fitting results of cumulative link mixed-effects model for blurred vision (disease cohort) | | | |
|---|---|---|---|
| AIC: 69100 BIC: 69200 Log-Likelihood: -34500 Number of Observations: 43483 Number of Users: 371 * | | | |
| **Random effect** | **Variance** | **Standard deviation** | |
| Individual (intercept) | 8.026 | 2.833 | |
| **Fixed effects** | **Estimate** | **Standard error** | **P value** |
| Follicular | -0.00358 | 0.0308 | 9.08E-01 |
| Early luteal | -0.157 | 0.0322 | 1.14E-06* |
| Late luteal | -0.14 | 0.0348 | 6.03E-05* |
| Premenstrual | -0.00234 | 0.0464 | 9.6E-01 |
| **Threshold coefficients** | **Estimate** | **Standard error** | **Z value** |
| 1 (Threshold 0 to 1) | -1.22 | 0.0986 | -12.4 |
| 2 (Threshold 1 to 2) | 1.95 | 0.0979 | 19.9 |
| 3 (Threshold 2 to 3) | 4.52 | 0.0992 | 45.5 |

| Table AA. The fitting results of cumulative link mixed-effects model for nerve pain (disease cohort) | | | |
|---|---|---|---|
| AIC: 60700 BIC:60800 Log-Likelihood: -30300 Number of Observations: 39730 Number of Users: 357 * | | | |
| **Random effect** | **Variance** | **Standard deviation** | |
| Individual (intercept) | 9.651 | 3.107 | |
| **Fixed effects** | **Estimate** | **Standard error** | **P value** |
| Follicular | -0.0165 | 0.033 | 6.17E-01 |
| Early luteal | -0.0614 | 0.0346 | 7.59E-02 |
| Late luteal | -0.0468 | 0.0373 | 2.1E-01 |
| Premenstrual | 0.0352 | 0.0496 | 4.78E-01 |
| **Threshold coefficients** | **Estimate** | **Standard error** | **Z value** |
| 1 (Threshold 0 to 1) | -1.02 | 0.169 | -6.04 |
| 2 (Threshold 1 to 2) | 2.04 | 0.169 | 12.1 |
| 3 (Threshold 2 to 3) | 4.84 | 0.171 | 28.3 |

| Table AB. The fitting results of cumulative link mixed-effects model for acid reflux (disease cohort) | | | |
|---|---|---|---|
| AIC: 52800 BIC: 52900 Log-Likelihood: -26400 Number of Observations: 35428 Number of Users: 329 * | | | |
| **Random effect** | **Variance** | **Standard deviation** | |
| Individual (intercept) | 4.691 | 2.166 | |
| **Fixed effects** | **Estimate** | **Standard error** | **P value** |
| Follicular | 0.0318 | 0.0359 | 3.76E-01 |
| Early luteal | 0.191 | 0.0373 | 3.09E-07* |
| Late luteal | 0.22 | 0.0402 | 4.32E-08* |
| Premenstrual | 0.101 | 0.054 | 6.19E-02 |
| **Threshold coefficients** | **Estimate** | **Standard error** | **Z value** |
| 1 (Threshold 0 to 1) | 0.611 | 0.125 | 4.88 |
| 2 (Threshold 1 to 2) | 2.98 | 0.126 | 23.6 |
| 3 (Threshold 2 to 3) | 5.18 | 0.131 | 39.6 |

| Table AC. The fitting results of cumulative link mixed-effects model for allergies (disease cohort) | | | |
|---|---|---|---|
| AIC: 50800 BIC: 50900 Log-Likelihood: -25400 Number of Observations: 31178 Number of Users: 283. | | | |
| **Random effect** | **Variance** | **Standard deviation** | |
| Individual (intercept) | 5.573 | 2.361 | |
| **Fixed effects** | **Estimate** | **Standard error** | **P value** |
| Follicular | 0.0176 | 0.0363 | 6.29E-01 |
| Early luteal | 0.037 | 0.0378 | 3.28E-01 |
| Late luteal | 0.192 | 0.0405 | 2.15E-06* |
| Premenstrual | 0.184 | 0.0536 | 6.13E-04* |
| **Threshold coefficients** | **Estimate** | **Standard error** | **Z value** |
| 1 (Threshold 0 to 1) | -0.411 | 0.145 | -2.83 |
| 2 (Threshold 1 to 2) | 2.26 | 0.146 | 15.5 |
| 3 (Threshold 2 to 3) | 4.9 | 0.15 | 32.6 |

| Table AD. The fitting results of cumulative link mixed-effects model for cough (disease cohort) | | | |
|---|---|---|---|
| AIC: 50600 BIC: 50600 Log-Likelihood: -25300 Number of Observations: 31768 Number of Users: 283 * | | | |
| **Random effect** | **Variance** | **Standard deviation** | |
| Individual (intercept) | 3.963 | 1.991 | |
| **Fixed effects** | **Estimate** | **Standard error** | **P value** |
| Follicular | -0.0706 | 0.0366 | 5.33E-02 |
| Early luteal | -0.0401 | 0.0379 | 2.9E-01 |
| Late luteal | 0.123 | 0.0408 | 2.45E-03* |
| Premenstrual | 0.00491 | 0.0547 | 9.28E-01 |
| **Threshold coefficients** | **Estimate** | **Standard error** | **Z value** |
| 1 (Threshold 0 to 1) | 0.286 | 0.125 | 2.29 |
| 2 (Threshold 1 to 2) | 2.65 | 0.126 | 21 |
| 3 (Threshold 2 to 3) | 4.77 | 0.131 | 36.5 |

| **Table AE. The fitting results of cumulative link mixed-effects model for tremors (disease cohort)** | | | |
|---|---|---|---|
| AIC: 52700 BIC: 52700 Log-Likelihood: -26300 Number of Observations: 32482 Number of Users: 271 * | | | |
| **Random effect** | **Variance** | **Standard deviation** | |
| Individual (intercept) | 5.869 | 2.423 | |
| **Fixed effects** | **Estimate** | **Standard error** | **P value** |
| Follicular | -0.0261 | 0.0356 | 4.64E-01 |
| Early luteal | -0.0219 | 0.0372 | 5.55E-01 |
| Late luteal | 0.0383 | 0.0402 | 3.41E-01 |
| Premenstrual | 0.146 | 0.0535 | 6.25E-03* |
| **Threshold coefficients** | **Estimate** | **Standard error** | **Z value** |
| 1 (Threshold 0 to 1) | -0.323 | 0.152 | -2.12 |
| 2 (Threshold 1 to 2) | 2.35 | 0.153 | 15.4 |
| 3 (Threshold 2 to 3) | 4.8 | 0.156 | 30.8 |

| **Table AF. The fitting results of cumulative link mixed-effects model for numbness (disease cohort)** | | | |
|---|---|---|---|
| AIC: 39900 BIC:40000 Log-Likelihood: -20000 Number of Observations: 28831 Number of Users: 271 * | | | |
| **Random effect** | **Variance** | **Standard deviation** | |
| Individual (intercept) | 8.68 | 2.946 | |
| **Fixed effects** | **Estimate** | **Standard error** | **P value** |
| Follicular | 0.00966 | 0.04 | 8.09E-01 |
| Early luteal | 0.0435 | 0.0417 | 2.97E-01 |
| Late luteal | 0.0526 | 0.0451 | 2.44E-01 |
| Premenstrual | 0.0369 | 0.0602 | 5.4E-01 |
| **Threshold coefficients** | **Estimate** | **Standard error** | **Z value** |
| 1 (Threshold 0 to 1) | -0.107 | 0.187 | -0.571 |
| 2 (Threshold 1 to 2) | 3.19 | 0.188 | 16.9 |
| 3 (Threshold 2 to 3) | 5.93 | 0.193 | 30.7 |

| **Table AG. The fitting results of cumulative link mixed-effects model for fever (disease cohort)** | | | |
|---|---|---|---|
| AIC: 32900 BIC: 33000 Log-Likelihood: -16500 Number of Observations: 27378 Number of Users: 224 * | | | |
| **Random effect** | **Variance** | **Standard deviation** | |
| Individual (intercept) | 5.3 | 2.302 | |
| **Fixed effects** | **Estimate** | **Standard error** | **P value** |
| Follicular | -0.266 | 0.0457 | 5.54E-09* |
| Early luteal | -0.131 | 0.0474 | 5.57E-03* |
| Late luteal | 0.135 | 0.0503 | 7.1E-03* |
| Premenstrual | 0.141 | 0.0665 | 3.42E-02* |
| **Threshold coefficients** | **Estimate** | **Standard error** | **Z value** |
| 1 (Threshold 0 to 1) | 1.05 | 0.162 | 6.5 |
| 2 (Threshold 1 to 2) | 3.46 | 0.164 | 21.1 |
| 3 (Threshold 2 to 3) | 5.79 | 0.171 | 33.9 |

| Table AH. The fitting results of cumulative link mixed-effects model for derealization (disease cohort) | | | |
|---|---|---|---|
| AIC: 30000 BIC: 30100 Log-Likelihood: -15000 Number of Observations: 19621 Number of Users: 201 * |  |  |  |
| **Random effect** | **Variance** | **Standard deviation** | |
| Individual (intercept) | 9.449 | 3.074 | |
| **Fixed effects** | **Estimate** | **Standard error** | **P value** |
| Follicular | 0.0315 | 0.047 | 5.04E-01 |
| Early luteal | -0.0577 | 0.0492 | 2.41E-01 |
| Late luteal | -0.107 | 0.0533 | 4.42E-02* |
| Premenstrual | 0.0222 | 0.0714 | 7.56E-01 |
| **Threshold coefficients** | **Estimate** | **Standard error** | **Z value** |
| 1 (Threshold 0 to 1) | -0.0489 | 0.228 | -0.215 |
| 2 (Threshold 1 to 2) | 2.31 | 0.228 | 10.1 |
| 3 (Threshold 2 to 3) | 4.85 | 0.232 | 20.9 |

| Table AI. The fitting results of cumulative link mixed-effects model for altered smell (disease cohort) | | | |
|---|---|---|---|
| AIC: 15700 BIC: 15700 Log-Likelihood: -7820 Number of Observations: 14551 Number of Users: 147 * |  |  |  |
| **Random effect** | **Variance** | **Standard deviation** | |
| Individual (intercept) | 15.43 | 3.928 | |
| **Fixed effects** | **Estimate** | **Standard error** | **P value** |
| Follicular | 0.0306 | 0.0639 | 6.32E-01 |
| Early luteal | 0.0742 | 0.0663 | 2.63E-01 |
| Late luteal | 0.0113 | 0.0719 | 8.75E-01 |
| Premenstrual | 0.0789 | 0.0961 | 4.11E-01 |
| **Threshold coefficients** | **Estimate** | **Standard error** | **Z value** |
| 1 (Threshold 0 to 1) | 0.841 | 0.341 | 2.47 |
| 2 (Threshold 1 to 2) | 4.45 | 0.343 | 12.9 |
| 3 (Threshold 2 to 3) | 7.73 | 0.355 | 21.8 |

| Table AJ. The fitting results of cumulative link mixed-effects model for altered taste (disease cohort) | | | |
|---|---|---|---|
| AIC: 13800 BIC: 13800 Log-Likelihood: -6880 Number of Observations: 13143 Number of Users: 135 * |  |  |  |
| **Random effect** | **Variance** | **Standard deviation** | |
| Individual (intercept) | 17.42 | 4.173 | |
| **Fixed effects** | **Estimate** | **Standard error** | **P value** |
| Follicular | 0.0413 | 0.0684 | 5.46E-01 |
| Early luteal | 0.0141 | 0.0712 | 8.43E-01 |
| Late luteal | 0.163 | 0.0765 | 3.36E-02* |
| Premenstrual | 0.102 | 0.103 | 3.2E-01 |
| **Threshold coefficients** | **Estimate** | **Standard error** | **Z value** |
| 1 (Threshold 0 to 1) | 0.924 | 0.378 | 2.44 |
| 2 (Threshold 1 to 2) | 4.48 | 0.381 | 11.7 |
| 3 (Threshold 2 to 3) | 7.73 | 0.39 | 19.8 |

**Table S7: The fitting results of binomial mixed-effects models for individual symptoms (that overlap exactly with the symptoms tracked in the disease cohort) in the population cohort**

| Table A. The fitting results of binomial link mixed-effects model for fatigue (population cohort) | | | |
|---|---|---|---|
| AIC: 17800 BIC: 17800 Log-Likelihood: -8890 Number of Observations: 29377 Number of Users: 377 * = | | | |
| **Random effect** | **Variance** | **Standard deviation** | |
| Individual (intercept) | -3.06 | 0.0943 | |
| **Fixed effects** | **Estimate** | **Standard error** | **P value** |
| Follicular | 0.3 | 0.0544 | **<0.001*** |
| Early luteal | 0.449 | 0.058 | **<0.001*** |
| Late luteal | 0.716 | 0.0614 | **<0.001*** |
| Premenstrual | 0.664 | 0.0808 | **<0.001*** |

| Table B. The fitting results of binomial mixed-effects model for anxiety (population cohort) | | | |
|---|---|---|---|
| AIC: 14500 BIC: 14500 Log-Likelihood: -7230 Number of Observations: 29377 Number of Users: 366 * = P | | | |
| **Random effect** | **Variance** | **Standard deviation** | |
| Individual (intercept) | -3.38 | 0.000264 | |
| **Fixed effects** | **Estimate** | **Standard error** | **P value** |
| Follicular | 0.407 | 0.000264 | **<0.001*** |
| Early luteal | 0.309 | 0.000264 | **<0.001*** |
| Late luteal | 0.661 | 0.000264 | **<0.001*** |
| Premenstrual | 0.843 | 0.000264 | **<0.001*** |

| Table C. The fitting results of binomial link mixed-effects model for brain fog (population cohort) | | | |
|---|---|---|---|
| AIC: 9420 BIC: 9470 Log-Likelihood: -4700 Number of Observations: 29377 Number of Users: 290 * = P | | | |
| **Random effect** | **Variance** | **Standard deviation** | |
| Individual (intercept) | -3.98 | 0.103 | |
| **Fixed effects** | **Estimate** | **Standard error** | **P value** |
| Follicular | -0.0487 | 0.0862 | 0.572 |
| Early luteal | 0.00573 | 0.0897 | 0.949 |
| Late luteal | 0.249 | 0.0924 | **0.007*** |
| Premenstrual | 0.521 | 0.113 | **<0.001*** |

| Table D. The fitting results of binomial link mixed-effects model for diarrhoea (population cohort) | | | |
|---|---|---|---|
| AIC: 9640 BIC: 9690 Log-Likelihood: -4820 Number of Observations: 29377 Number of Users: 274 * = P | | | |
| **Random effect** | **Variance** | **Standard deviation** | |
| Individual (intercept) | -5.11 | 0.123 | |
| **Fixed effects** | **Estimate** | **Standard error** | **P value** |
| Follicular | 1.72 | 0.0896 | **<0.001*** |
| Early luteal | 1.8 | 0.0936 | **<0.001*** |
| Late luteal | 1.41 | 0.105 | **<0.001*** |
| Premenstrual | 1.56 | 0.126 | **<0.001*** |

| Table E. The fitting results of binomial link mixed-effects model for loss of appetite (population cohort) | | |
|---|---|---|
| AIC: 8580 BIC: 8630 Log-Likelihood: -4280 Number of Observations: 29377 Number of Users: 229 * = P | | |
| **Random effect** | **Variance** | **Standard deviation** |
| Individual (intercept) | -4.65 | 0.141 |

| **Fixed effects** | **Estimate** | **Standard error** | **P value** |
|---|---|---|---|
| Follicular | 0.319 | 0.0815 | **<0.001*** |
| Early luteal | 0.113 | 0.0914 | 0.217 |
| Late luteal | -0.0999 | 0.107 | 0.350 |
| Premenstrual | 0.236 | 0.13 | 0.069 |

| **Table F. The fitting results of binomial link mixed-effects model for depression (population cohort)** | | | |
|---|---|---|---|
| AIC: 6210 BIC: 6260 Log-Likelihood: -91900 Number of Observations: 29377 Number of Users: 223 * = P | | | |
| **Random effect** | **Variance** | **Standard deviation** | |
| Individual (intercept) | -4.9 | 0.128 | |
| **Fixed effects** | **Estimate** | **Standard error** | **P value** |
| Follicular | 0.137 | 0.115 | 0.232 |
| Early luteal | 0.309 | 0.117 | **0.008*** |
| Late luteal | 0.748 | 0.113 | **<0.001*** |
| Premenstrual | 1.06 | 0.133 | **<0.001*** |

| **Table G. The fitting results of binomial link mixed-effects model for nausea (population cohort)** | | | |
|---|---|---|---|
| AIC: 6860 BIC: 6910 Log-Likelihood: -3420 Number of Observations: 29377 Number of Users: 223 * = P | | | |
| **Random effect** | **Variance** | **Standard deviation** | |
| Individual (intercept) | -5.08 | 0.142 | |
| **Fixed effects** | **Estimate** | **Standard error** | **P value** |
| Follicular | 0.29 | 0.104 | **0.005*** |
| Early luteal | 0.69 | 0.102 | **<0.001*** |
| Late luteal | 0.84 | 0.11 | **<0.001*** |
| Premenstrual | 0.857 | 0.141 | **<0.001*** |

| **Table H. The fitting results of binomial link mixed-effects model for constipation (population cohort)** | | | |
|---|---|---|---|
| AIC: 5150 BIC: 5200 Log-Likelihood: -2570 Number of Observations: 29377 Number of Users: 150 * = P | | | |
| **Random effect** | **Variance** | **Standard deviation** | |
| Individual (intercept) | -6.47 | 0.267 | |
| **Fixed effects** | **Estimate** | **Standard error** | **P value** |
| Follicular | 0.414 | 0.119 | **<0.001*** |
| Early luteal | 0.685 | 0.119 | **<0.001*** |
| Late luteal | 0.898 | 0.126 | **<0.001*** |
| Premenstrual | 0.765 | 0.167 | **<0.001*** |

| **Table I. The fitting results of binomial link mixed-effects model for blurred vision (population cohort)** | | | |
|---|---|---|---|
| AIC: 2770 BIC: 2820 Log-Likelihood: -1380 Number of Observations: 29377 Number of Users: 96 * = P | | | |
| **Random effect** | **Variance** | **Standard deviation** | |
| Individual (intercept) | -8.16 | 0.551 | |
| **Fixed effects** | **Estimate** | **Standard error** | **P value** |
| Follicular | 0.229 | 0.166 | 0.168 |
| Early luteal | 0.367 | 0.174 | **0.035*** |
| Late luteal | 0.365 | 0.188 | 0.053 |
| Premenstrual | 0.0373 | 0.283 | 0.895 |

**Supplementary Methods**

**Data source and collection – how users track symptoms and how survey was conducted (disease cohort)**
Users select their commonly experienced symptoms from the following categories: general (fatigue, fever, allergies, tremors), brain (headache, brain fog, anxiety, dizziness, depression, derealisation, memory issues, lightheadedness), heart and lungs (palpitations, cough, shortness of breath), pain (sore throat, nerve pain, migraine, chest pain, joint pain, stomach pain), muscles (muscle aches, muscle weakness), sensory (altered smell, altered taste, pins & needles, tinnitus, light sensitivity, numbness, blurred vision, noise sensitivity) and gastrointestinal (diarrhoea, nausea, acid reflux, constipation, lack of appetite). Users can also input their own custom symptoms.

Each day, users are prompted to rate their commonly experienced symptoms on a 4-point Likert scale [None, Mild, Moderate, Severe] and whether they are currently experiencing a crash (the sudden and severe worsening of symptoms following physical, mental or emotional exertion[2]) [No, Yes]. Users are also asked whether they are currently having their menstrual period [No, Yes].

Upon consent to participate in our study, users were asked the following 5 survey questions: (1) Are you using any hormonal contraception? [No, Combined pill, Progestogen-only pill, Intrauterine system (IUS), Contraceptive Injection, Contraceptive implant, Contraceptive patch]; (2) Are you currently pregnant [No, Yes]; (3) Are you currently breastfeeding [No, Yes]; (4) How many times have you been pregnant? [0, 1, 2, 3 or more]; (5) How many times have you given birth? [0, 1, 2, 3 or more].

**Data source and collection – how users track symptoms and how the survey was conducted (population cohort)**
Upon downloading the Hertility app, users are asked to complete an online health assessment. This includes age, gender identity, ethnicity, cycle regularity, current form of contraception, and whether they have been pregnant before. Users can then track the following symptoms:

Flow: Heaviness [Spotting, Light, Moderate, Heavy]; Colour [Bright red, Brown/dark red, Light pink, Grey, Black, Orange]; Jelly-like blood clots [Small, Medium, Large]

Mood: [Anxiety, Balanced, Brain fog, Comfortable, Confused, Confident, Depressed, Empowered, Energised, Frisky, Irritable, Low energy, Low mood, Mood swings, Obsessive thoughts, Patient, Reflective, Relaxed, Resilient, Sad, Self-critical]

Pain: Pelvic pain [Uncomfortable, Intense, Unbearable]; Period cramps [Uncomfortable, Intense, Unbearable]; Backache [Uncomfortable, Intense, Unbearable]; External pain during intercourse; Internal pain during intercourse; Bleeding after intercourse; Ovulation pain; Leg pain

Sleep and temperature: Insomnia; Fatigue; Night shift work; Night sweats; Hot flushes; Feeling cold; Basal temperature

Head & Skin: Excessive body or facial hair; Hair thinning or loss; Blurry vision; Headaches [Uncomfortable, Intense, Unbearable]; Migraine [Uncomfortable, Intense, Unbearable]; Dark skin patches; Acne [Almost clear, Mild, Moderate, Severe]

Breasts: Tender breasts; Swollen breasts; Nipple discharge; Itchy nipples

Vulva & Vagina: Itchy vulva; Vaginal dryness; Vaginal discharge [Watery, Creamy, Sticky, Egg white, Clumpy white, White, Yellow, Pink, Red, Brown, Green, Grey, Black]

Appetite & Digestion: Increased hunger; Cravings; Loss of appetite; Bloating; Nausea or vomiting; Painful peeing; Leaking pee; Peeing every 2 hours; Continuous need to pee; Inability to fully empty your bladder; Can barely hold your pee; Painful bowel movements; Poo [Hard lumps, Lumpy sausage-shaped, Cracked sausage-shaped, Smooth sausage-shaped, Clear-cut soft blobs, Mushy, Watery or liquid]

Activity: Sexual intercourse [No intercourse, Protected intercourse, Unprotected intercourse]; Travelling; Stress; Sick; Change in medications; Exercise [Low, Moderate, High]

Alcohol & Drugs: Hungover; Alcohol; Cigarette; Vaping; Recreational drugs

**Data cleaning (population cohort)**

As with the disease cohort analysis, cleaning and analysis of the Hertility data was conducted using R version 4.5.1 (2025-06-13) in RStudio version 2025.5.1.513. [3] User-level demographic data from the online health assessment was matched with daily app-based symptom observation data, and only users appearing in both the datasets were retained. Observations recorded before the Visible study period start date (7 September 2022) were excluded. Users outside the age criteria were excluded and age was categorised into <30, 30-39, >40. Contraception type was categorised into no hormonal contraception, progestin-only, combined oestrogen and progestin. Any users that did not track any 'flow' symptoms were excluded. When a flow symptom was tracked = on period, when a flow symptom was not tracked = not on period. Period start date and menstrual phases were assigned using the method described below. Cycles outside range of 24–38 days were excluded. This is visualised in **Figure S2**. To ensure consistency in tracking across the cycle, only users with at least one non-flow symptom report in every phase were retained (see **Figure S3 and S4** for before and after this filtering stage for number of observations reported in each menstrual phase).

**Demographics of user groups at each stage of data cleaning (disease cohort)**

Corresponds to Table S1. 1932 users filled out the survey, were within the age range of 18-45, had long COVID and/or ME/CFS, and were not breastfeeding and/or pregnant. The 1932 users were filtered down to 1765 users by removing any users who did not track their periods or report any bleeding days. The 1765 users were then filtered down to 1109 users by excluding cycles that had >2 consecutive days of missing menstrual data. The 1109 users were then filtered down to 948 users by excluding cycles that were abnormally short (<24 days) or long (>38 days). At each stage of filtering, we summarised participant characteristics, including disease group, contraception use, and nulligravida status, as well as age, mean daily symptom score, and the percentage of days with crash. Categorical variables were compared using $\chi^2$ tests, and continuous variables were compared using Kruskal–Wallis tests.

**Menstrual cycle phase coding (both cohorts)**

A menstrual cycle was counted from the first day of one menstrual period to the first day of the next. The first day of a period was assigned based on the International Federation of Gynecology and Obstetrics (FIGO) definition of a normal period[4] where the day fulfilled all the following criteria: user reported being on their period; user did not report being on their period the previous 5 days; user either reported being on their period or did not make a report on the following two days; user did not report being on their period 9 and 10 days after. More than 8 days of bleeding indicates an abnormally long period, or continuous spotting associated with progestin-only contraception; fewer than two days of bleeding indicates spotting rather than a period. The period end date was assigned when the day fulfilled all the following criteria: user reported being on their period; user reported being on their period for at least the previous 2 days; user did not report being on their period the following day. Menstrual cycles that contained >2 consecutive days of missing menstrual tracking data (for the disease cohort) or were outside of the normal length of a cycle (24-38 days, as defined by FIGO[4]) were removed from the dataset. These cycles are either abnormally short or long, or represent a failure to report bleeding days. Exclusion of users and cycles is summarised in **Figure 1B** for the disease cohort and in **Figure S2** for the population cohort.

For all remaining cycles, any missing menstrual tracking data was imputed ('on period' or 'not on period') so that menstrual cycle phases could be assigned. In the disease cohort, a total of 7303 days (5.68% of all days) had imputed menstrual cycle data, but many fewer than this are at risk of being misassigned because of our approach of assigning cycles before imputing missing data, and of imputing no more than two consecutive days of data. Menstrual data was imputed from the previous day with data, with the following exception: missing data on the 9$^{th}$ or 10$^{th}$ day of the cycle was imputed as 'not on period', assuming duration of menses does not exceed the normal 8 days. [4] Therefore, the only days at risk of being misassigned are days with no report that immediately follow a reported bleeding day. There is a total of 1119 such days in the dataset, 0.87% of all days. This approach differs for the population cohort, in which users report only days when they are menstruating; consequently, any day without a reported period is assumed to be a non-menstrual day.

A menstrual cycle phase was assigned to each day of all included cycles according to the decision tree outlined in **Figure 1C.** This follows the assumption that the follicular phase is variable, and the luteal phase consistently accounts for the final 14 days of each cycle.[5] The luteal phase was further divided into the early luteal (14-8 days before the first day of period (D1), late luteal (7-3 days before D1) and premenstrual phase (2 days before D1).

**Demographics of study population (both cohorts)**
The age distributions of the disease cohort and the population cohort are presented in **Figures S5** and **S6**, respectively. Demographic characteristics of the disease cohort are summarised in Table 1 and further stratified by disease type (**Table S2**) and contraception type (**Table S3**). Differences in age were assessed using the Kruskal–Wallis rank-sum test, while differences in all other demographic variables were assessed using Pearson's $\chi^2$ tests. Age distributions across contraception types are shown in **Figure S7**. Associations between age and disease type were evaluated using Kruskal–Wallis tests. Where overall tests were significant, post hoc pairwise comparisons were performed using Dunn's tests, with Bonferroni adjustment applied to control for type I error. Demographic characteristics of the population cohort are summarised in **Table S4**.

**Descriptive statistics and statistical testing (disease cohort)**
For overall study population characteristics, categorical variables were presented as numbers and percentages. The distribution of the mean overall symptom score for each individual across all reports, is presented as a histogram is not normally distributed (Shapiro-Wilk test) (**Figure S8**). Unless otherwise specified, all statistical tests were run using the inbuilt *stats* R package, [6] and all plots were created using the *ggplot2* package. [7] The Friedman test was used to test the significance between mean symptom score and menstrual phase, due to the repeated measures nature of the data. The Mann-Whitney U-Test was used to test the significance between gravidity and mean symptom score, due to the binary nature of the variable. Unpaired Kruskal-Wallis tests were used to test significance for all other variables. P-values were adjusted using the Bonferroni method to control for Type 1 error, using the *FSA* package.[8]

**Prevalence and clustering of symptoms (disease cohort, supplementary only)**
The distribution of users tracking symptoms over the study period was grouped into quarters-post-joining and visualised using a line graph (**Figure S9**). The number and percentage of the study population (n=948) tracking each of the standard 36 symptoms at least once was calculated, and the distribution of the total number of days each symptom was tracked was visualised (**Figure S10**). A Jaccard similarity heatmap and hierarchical clustering was used to identify clusters of symptoms commonly co-reported by users (**Figure S11,12**). For each of the 594 users, for all 36 symptoms, it was noted whether the user had tracked the symptom (1) or had never tracked the symptom (0), and results arranged in a 594 x 36 matrix. Then Jaccard similarities for each pair of symptoms (*A* and *B*) was computed by dividing the number of users tracking at least one of the symptoms by the number of users tracking both symptoms:

$$Jaccard\ similarity\ (A,B) = \frac{|A \cap B|}{|A \cup B|} \qquad Jaccard\ distance\ (A,B) = 1 - \frac{|A \cap B|}{|A \cup B|}$$

Jaccard distances were used for hierarchical clustering using the complete linkage method, where the distance between clusters is determined by the maximum distance between any pair of points from the two clusters. The R package *pheatmap* was used to create all heatmaps.

**Effect of menstrual cycle phase on individual symptoms (both cohorts, supplementary only)**
For each symptom in the disease cohort, a contingency table was constructed to show the total number of reported days by symptom severity [None (0), Mild (1), Moderate (2), Severe (3)] and assigned menstrual cycle phase [Menstrual, Follicular, Early Luteal, Late Luteal, Premenstrual]. For each aspect of health tracked by the population cohort that overlapped exactly with a symptom that was tracked in the disease cohort, a contingency table was constructed to show the total number of reported days by symptom presence [Not present, Present] and assigned menstrual cycle phase [Menstrual, Follicular, Early Luteal, Late Luteal, Premenstrual]. The *spineplot* function from the inbuilt R *graphics* package[6] was used to create spineplots to visualise data for each symptom. For each phase of the menstrual cycle, symptom spineplots show the proportion of each symptom severity rating/symptom presence, with the width of the bars indicating the total number of counts in that bar. Spineplots are shown in **Figure S13** for the disease cohort and **Figure S14** for the population cohort.

**Mixed effects modelling – crashes**

The crashes model was fitted using the *glmer* function from the *lme4* package[9], which also uses maximum likelihood estimation with Laplace approximation to integrate over random effects. As the outcome variable is "crash" or "no crash", we use a binomial model.

The model AIC was 81256·9, the BIC was 81428·4, the total number of observations analysed was 101953, from a total of 948 users. The model equation was as follows:

$$S \sim \text{Bernoulli}\left(u_{ij}, \theta\right) \qquad c_j \sim \text{N}\,(0, \text{G})$$
$$\log\left(u_{ij}\right) = \beta_0 + \beta_1 \cdot Menstrual\ phase + \beta_2 \cdot Contraception\ type + \beta_3 \cdot Disease\ type$$
$$+ \beta_4 \cdot Age\ category + \beta_5 \cdot Nulligravidity + \beta_6 \cdot Quarters\ post\ joining + c_j$$

Contraception was categorised into no hormonal contraception, combined hormonal contraception (combined pill, contraceptive patch) and progestin-only contraception (intrauterine system, progestogen-only pill, contraceptive implant), and age was categorised into under 30, 30-39 and 40 or over, to ensure there were similar numbers in each category. Time was categorised as quarters post-joining, with the join date being the date of the first report submitted by a given individual and a quarter being defined as three months.

Where:

- $S$ is the response variable indicating crash occurrence
- $u_{ij}$ is the expected crash occurrence, for date $i$ and individual $j$
- θ is the dispersion parameter of the Bernoulli distribution
- $\beta_0$ is the intercept and $\beta_1$-$\beta_6$ are the coefficients for the fixed effects
- $c_j$ is the random intercept for individual $j$
- $G$ is the variance of the random intercept for individual $j$

**Mixed effects modelling – overall symptom score**

The overall symptom score model was fitted using the *glmer.nb* function from the *lme4* package[9] initialised via *theta.ml* from the *MASS* package[10] and employing maximum likelihood estimation with Laplace approximation to integrate over random effects. The negative binomial mixed-effects model assumes the overall symptom score is a non-negative integer and accounts for the overdispersion in the data (present in most count data, where the variance is greater than the mean[11]) and is appropriate for our data (**Figure S8A**).

The model AIC was 689187·3, the BIC was 689371·6, the total number of observations analysed was 120916, from a total of 948 users. The model equation was as follows:

$$S \sim \text{Negative Binomial}\left(u_{ij}, \theta\right) \qquad c_j \sim \text{N}\,(0, \text{G})$$
$$\log\left(u_{ij}\right) = \beta_0 + \beta_1 \cdot Menstrual\ phase + \beta_2 \cdot Contraception\ type + \beta_3 \cdot Disease\ type$$
$$+ \beta_4 \cdot Age\ category + \beta_5 \cdot Nulligravidity + \beta_6 \cdot Quarters\ post\ joining + c_j$$

Where the variables represent the same components as in the crash occurrence model, with $S$ now representing symptom score.

**Mixed effects modelling – fatigue, brain fog and headache**

The top 3 most tracked individual symptom models were fitted on the disease cohort data using the *clmm* function from the *ordinal* package.[12] These models were cumulative link mixed-effects models (CLMMs), appropriate for ordinal data (individual symptom scores ranged from 0 to 3). The models employed a logit link function and maximum likelihood estimation with Laplace approximation to account for random effects. The max iterations were set to 40,000, and the maximum absolute gradient of the inner optimisation was set to 1e-4. Fixed effects are the same as the crash and overall symptom score models except quarters post-joining was not included as this measure wasn't able to be obtained for the population cohort. The model equations were as follows:

$$S \sim \text{Ordinal}\left(u_{ij}, \tau\right) \qquad c_j \sim \text{N}\,(0, \text{G})$$
$$\log\left(u_{ij}\right) = \beta_0 + \beta_1 \cdot Menstrual\ phase + \beta_2 \cdot Contraception\ type + \beta_3 \cdot Disease\ type$$
$$+ \beta_4 \cdot Age\ category + \beta_5 \cdot Nulligravidity + c_j$$

Where:

- $S$ is the ordinal response variable indicating the score of an individual symptom (0, 1, 2, or 3)

- $u_{ij}$ is the latent continuous variable used to model the ordinal response via cumulative thresholds, for date $i$ and individual $j$
- τ represents the threshold coefficients that separate the ordinal categories
- $\beta_0$ is the intercept and $\beta_{1-5}$ are the fixed effect coefficients
- $c_j$ is the random intercept for individual $j$
- $G$ is the variance of the random intercept for individual $j$

Fatigue, brain fog and headache comparator models were fitted on the population cohort data using the *glmer* function from the *lme4* package[9], which also uses maximum likelihood estimation with Laplace approximation to integrate over random effects. As the outcome variable is "present" or "not present", we used binomial models. The model equation was as follows:

$$S \sim \text{Bernoulli}\left(u_{ij}, \theta\right) \qquad c_j \sim \text{N}\,(0, \text{G})$$
$$\log\left(u_{ij}\right) = \beta_0 + \beta_1 \cdot Menstrual\ phase + \beta_2 \cdot Contraception\ type + \beta_3 \cdot Disease\ type + \beta_4 \cdot Age\ category + \beta_5 \cdot Nulligravidity + c_j$$

Where:

- $S$ is the response variable indicating symptom presence
- $u_{ij}$ is the expected symptom presence, for date $i$ and individual $j$
- θ is the dispersion parameter of the Bernoulli distribution
- $\beta_0$ is the intercept and $\beta_1$-$\beta_5$ are the coefficients for the fixed effects
- $c_j$ is the random intercept for individual $j$
- $G$ is the variance of the random intercept for individual $j$

**Mixed effects modelling – fatigue, brain fog and headaches joint dataset models (supplementary only)**
To enable joint modelling of symptoms across cohorts, symptom data from the Visible and Hertility datasets were combined using a full join prior to analysis, for each of the top 3 symptoms. A new variable 'cohort type' was created [disease cohort, population cohort]. An interaction term for cohort type and menstrual phase was included to estimate how much the menstrual phase effect changes in the disease cohort relative to the population cohort. In the population dataset, each health variable was originally recorded as binary and was rescaled to match the 0–3 ordinal scale used in the visible dataset by recoding presence of fatigue as 3 and absence as 0. Symptoms were then treated as an ordered factor with levels 0–3. The joint dataset models were fitted using the *clmm* function from the *ordinal* package.[12] Cumulative link mixed-effects models (CLMMs) were used as they are appropriate for ordinal data. The models employed a logit link function and maximum likelihood estimation with Laplace approximation to account for random effects. The max iterations were set to 40,000, and the maximum absolute gradient of the inner optimisation was set to 1e-4. The model equations were as follows:

$$S \sim \text{Ordinal}\left(u_{ij}, \tau\right) \qquad c_j \sim \text{N}\,(0, \text{G})$$
$$\log\left(u_{ij}\right) = \beta_0 + \beta_1 \cdot (Cohort\ type * Menstrual\ phase) + \beta_2 \cdot Contraception\ type + + \beta_3 \cdot Age\ category + \beta_4 \cdot Nulligravidity + c_j$$

Where:

- $S$ is the ordinal response variable indicating the score of an individual symptom (0, 1, 2, or 3)
- $u_{ij}$ is the latent continuous variable used to model the ordinal response via cumulative thresholds, for date $i$ and individual $j$
- τ represents the threshold coefficients that separate the ordinal categories
- $\beta_0$ is the intercept and $\beta_{1-4}$ are the fixed effect coefficients
- $c_j$ is the random intercept for individual $j$
- $G$ is the variance of the random intercept for individual $j$

**Mixed effects modelling – 36 individual symptoms in disease cohort (supplementary only)**
The 36 individual symptom models were fitted using the *clmm* function from the *ordinal* package.[12] For the overall symptom score above, we could treat $u_{ij}$ it as a count variable; however, for the individual models, we needed to treat the data as ordinal. These models were cumulative link mixed-effects models (CLMMs), appropriate for ordinal data (individual symptom scores ranged from 0 to 3). The models employed a logit link function and maximum likelihood estimation with Laplace approximation to account for random effects. The simplest form of the model was used, which was to have menstrual phase as a fixed effect and 'individual' as a random effect. All symptoms were run with the default optimiser *nlminb*, except "lack of appetite" and "pins and needles" which were run with *ucminf*. The max iterations were set to 40,000, and the maximum absolute gradient of the inner optimisation was set to 1e-4. The model equations were as follows:

$$S \sim \text{Ordinal}\left(u_{ij}, \tau\right) \qquad c_j \sim \text{N}\left(0, \text{G}\right)$$
$$\log\left(u_{ij}\right) = \beta_0 + \beta_1 \cdot Menstrual\ phase + c_j$$

Where:

- $S$ is the ordinal response variable indicating the score of an individual symptom (0, 1, 2, or 3)
- $u_{ij}$ is the latent continuous variable used to model the ordinal response via cumulative thresholds, for date $i$ and individual $j$
- $\tau$ represents the threshold coefficients that separate the ordinal categories
- $\beta_0$ is the intercept and $\beta_1$ is the coefficient for the fixed effect 'menstrual phase'
- $c_j$ is the random intercept for individual $j$
- $G$ is the variance of the random intercept for individual $j$

**Mixed effects modelling – 9 individual symptoms in population cohort that overlap with disease cohort symptoms (supplementary only)**
There were 9 aspects of health that were tracked by the population cohort that were tracked as symptoms by the disease cohort. These 9 'symptoms' were fitted on the population cohort data using the *glmer* function from the *lme4* package[9], which also uses maximum likelihood estimation with Laplace approximation to integrate over random effects. As the outcome variable is "present" or "not present", we used binomial models. As with the 36 individual disease cohort models, the simplest form of the model was used, which was to have menstrual phase as a fixed effect and 'individual' as a random effect.

The model equations was as follows:

$$S \sim \text{Binomial}\left(u_{ij}, \theta\right) \qquad c_j \sim \text{N}\left(0, \text{G}\right)$$
$$\log\left(u_{ij}\right) = \beta_0 + \beta_1 \cdot Menstrual\ phase + c_j$$

Where the variables represent the same components as in the symptom score model, with $S$ now representing symptom occurrence.

**References for Supplementary Information**

1 Scientific Image and Illustration Software | BioRender. https://www.biorender.com/ (accessed Aug 8, 2024).

2 Parker M, Sawant HB, Flannery T, *et al.* Effect of using a structured pacing protocol on post-exertional symptom exacerbation and health status in a longitudinal cohort with the post-COVID-19 syndrome. *J Med Virol* 2023; 95. DOI:10.1002/jmv.28373.

3 Posit team. RStudio: Integrated Development Environment for R. 2024.

4 Munro MG, Critchley HOD, Fraser IS, *et al.* The two FIGO systems for normal and abnormal uterine bleeding symptoms and classification of causes of abnormal uterine bleeding in the reproductive years: 2018 revisions. *International Journal of Gynecology and Obstetrics* 2018; 143. DOI:10.1002/ijgo.12666.

5 Reed BG, Carr BR. The Normal Menstrual Cycle and the Control of Ovulation. 2000.

6 R Core Team. R: A Language and Environment for Statistical Computing_. R Foundation for Statistical Computing, Vienna, Austria. 2023. https://www.R-project.org/ (accessed Aug 2, 2024).

7 Wickham H, Chang W, Henry L, Lin-Pedersen T. Package 'ggplot2' Elegant Graphics for Data Analysis. 2016.

8 Ogle DH, Doll JC, Wheeler AP, Dinno A. FSA: Simple Fisheries Stock Assessment Methods. 2025. https://CRAN.R-project.org/package=FSA (accessed Jan 22, 2025).

9 Bates D, Mächler M, Bolker BM, Walker SC. Fitting linear mixed-effects models using lme4. *J Stat Softw* 2015; 67. DOI:10.18637/jss.v067.i01.

10 Venables WN, Ripley BD. Modern Applied Statistics with S Fourth edition by. 2002.

11 Ver Hoef JM, Boveng PL. Quasi-poisson vs. negative binomial regression: How should we model overdispersed count data? *Ecology* 2007; 88. DOI:10.1890/07-0043.1.

12 Christensen RHB. ordinal---Regression Models for Ordinal Data. 2023. https://CRAN.R-project.org/package=ordinal (accessed Jan 22, 2025).
